# Global prediction of unreported SARS-CoV2 infection from observed COVID-19 cases

**DOI:** 10.1101/2020.04.29.20083485

**Authors:** Carson C. Chow, Joshua C. Chang, Richard C. Gerkin, Shashaank Vattikuti

## Abstract

Estimation of infectiousness and fatality of the SARS-CoV-2 virus in the COVID-19 global pandemic is complicated by ascertainment bias resulting from incomplete and non-representative samples of infected individuals. We developed a strategy for overcoming this bias to obtain more plausible estimates of the true values of key epidemiological variables. We fit mechanistic Bayesian latent-variable SIR models to confirmed COVID-19 cases, deaths, and recoveries, for all regions (countries and US states) independently. Bayesian averaging over models, we find that the raw infection incidence rate underestimates the true rate by a factor, the case ascertainment ratio CAR_t_ that depends upon region and time. At the regional onset of COVID-19, the predicted global median was 13 infections unreported for each case confirmed (CAR_t_ = 0.07 C.I. (0.02, 0.4)). As the infection spread, the median CAR_t_ rose to 9 unreported cases for every one diagnosed as of April 15, 2020 (CAR_t_ = 0.1 C.I. (0.02, 0.5)). We also estimate that the median global initial reproduction number R_0_ is 3.3 (C.I (1.5, 8.3)) and the total infection fatality rate near the onset is 0.17% (C.I. (0.05%, 0.9%)). However the time-dependent reproduction number R_t_ and infection fatality rate as of April 15 were 1.2 (C.I. (0.6, 2.5)) and 0.8% (C.I. (0.2%,4%)), respectively. We find that there is great variability between country- and state-level values. Our estimates are consistent with recent serological estimates of cumulative infections for the state of New York, but inconsistent with claims that very large fractions of the population have already been infected in most other regions. For most regions, our estimates imply a great deal of uncertainty about the current state and trajectory of the epidemic.

## Introduction

Nearly 3 million confirmed cases of the novel coronavirus SARS-CoV-2 infection have been reported worldwide as of late-April, 2020 (“COVID-19 Map” 19). However, it is presumed that many infections remain unreported due to either mildness of symptoms or inadequate testing. Knowledge of the full extent of the COVID-19 pandemic is required to evaluate the effectiveness of mitigation strategies such as social distancing, which have a major economic and social cost.

In the absence of universal testing, a proportion of the infected either do not cross the diagnostic threshold for COVID-19 testing or are unable to acquire medical attention and thus remain undetected. Consequently, the fraction of infected who are reported as cases may be substantially less than 1. Estimating the total case ascertainment ratio (CAR_t_), defined as the ratio of the total number of confirmed cases to the total number of individuals infected with the novel coronavirus SARS-CoV-2 on day t, is thus important for constraining the initial reproduction number R_0_ of COVID-19, as well as recovery and fatality rates due to the disease.

Estimating the true magnitude and dynamics of the fractions of the population who are infected, susceptible, or recovered is a difficult and open problem. Serological and molecular diagnostic tests may not have perfect specificity and sensitivity (a substantial problem if the true positive rates are also low), may not be widely available, and may continue to suffer from ascertainment bias. Recent modeling work has attempted to estimate the true infected population, often relying on reported deaths since there is less ambiguity in the definition (Lourenco et al.; Flaxman et al.). Flaxman et. al 2020 focused on death data from the European Centre of Disease Control and suggest that there are orders of magnitude more infected than detected in confirmed cases. This claim was supported by a separate SIR model fitting cases, case recoveries, and case deaths that predicted a CAR of 1/63 for Italy (Calafiore et al.). Recent, preliminary, serological test results have been mixed. A study from Benavid et. al (Bendavid et al.) is consistent with this high number of undiagnosed cases. However a study of women admitted for delivery estimated that 1 in 8 cases are symptomatic (Sutton et al.), which is consistent with a recent press report by New York state based on sampling grocery store customers (New York Times). Debates continue on the adequacy of these tests.

Here, we develop a set of Bayesian, mechanistic, latent-variable, SIR models (Kermack and McKendrick; Wo and Ag, “Contributions to the Mathematical Theory of Epidemics--II. The Problem of Endemicity.1932”; Wo and Ag, “Contributions to the Mathematical Theory of Epidemics--III. Further Studies of the Problem of Endemicity. 1933”) that respect the uncertainty in the underlying data. We explore multiple specifications and find general consensus among the models. Our models try to account for the effects of mitigation (such as social distancing), the possibility that CAR_t_ (either through changes in definition or in detection, for example via ramp-up of testing) can vary in time, excess variability due to reporting and heterogeneity in the regional population, and incomplete quarantine of reported active cases. A similar model assumed complete quarantine on the basis of debilitating effects of illness or strict adherence to policy (Pedersen and Meneghini), which we relax to quantify the effectiveness of such actions. Using the model, we predict the total number of infected individuals from the confirmed numbers of cases, case recoveries, and case deaths. These three data time-series are sufficient to constrain time-varying estimates for CAR_t_ as well as for the fraction of unobserved infected individuals from which we can obtain the total reproduction number R_t_ and total Infection Fatality Ratio (IFR_t_) as functions of time.

## Methods

Our overall approach uses latent-variable Bayesian models with underlying dynamics given by a family of SIR models (Kermack and McKendrick). We implement our models in the Bayesian inference software language Stan (Carpenter et al.; Hartikainen et al.) to estimate the model statistics within each region independently, using data from The Johns Hopkins University Center for Systems Science and Engineering (JHU CSSE; Dong et al.) and from The COVID Tracking Project (“The COVID Tracking Project”). Collectively, these data consist of daily counts for the reported new cases, deaths, and recoveries (as well as key regional mitigation dates, where available (Flaxman et al.; Mervosh et al.)). We only consider regions that report all three of these quantities, which eliminates several US States and dates beyond April 15 for which the complete data was unavailable.

In the SIR models that we use to quantify the regional population dynamics of COVID-19, interactions between susceptible individuals (*S*) and infected individuals (*I*) induce an increase in *I* and a concomitant decrease in *S*. The unobserved members of *I* can either transition into a reported SARS-CoV-2 case, *C*, or become no longer infectious (either by recovering or dying). Confirmed cases *C* can then either recover (*R_C_*) or die (*D_C_*) and also infect more *S* at a rate that can differ from I. Mitigation efforts (including any social or policy changes aimed at reducing transmission rates) are modeled by a time dependence on the rate of infectious transmission. This rate changes from an initial value to a final value on a specified day with a 5 day transition period. We also include a similar time dependence in the rate of transition from *I* to *C* to account for changes in testing regimens, clinical classification, and awareness of infected individuals to seek medical attention. We consider six variations of the latent-variable SIR model of varying complexity. Besides the standard nonlinear SIR models, we also fit linear approximations that assume that S is approximately fixed (i.e. the epidemic is small enough that herd immunity is not significantly affecting transmission). The linear models have exponential growth with growth rates that can vary in time including becoming negative.

We tie each underlying SIR model to the available data using the generative probabilistic scheme shown in Figure 1. We assume a Negative Binomial likelihood, where the variance is a fitted parameter, for the observable data given the model predictions. This formulation accounts for the inherent variability of an underlying stochastic birth-death process (which usually obeys a Poisson distribution), disease progression effects, heterogeneous mixing of population clusters, and errors in classification and reporting inherent in the available data. We modeled prior probabilities for recovery and death rates using inverse gamma distributions derived from ref. (Verity et al.). For the other unknown distributions, we used weakly-informative priors for satisfying positivity and providing numerical and inferential stability. Priors for the day of mitigation application, taken from ref. (Flaxman et al.) and (Mervosh et al.), were used when available. We assessed models by balancing the likelihood of observing the data (given each model) against model complexity using a standard Bayesian model comparison metric such as Leave-One-Out cross validation (LOO) and WAIC (Vehtari et al.), to avoid overfitting. On the basis of this and other Bayesian metrics, we also report model-averaged statistics that take into account model specification uncertainty (Yao et al.).

**Figure 1:**
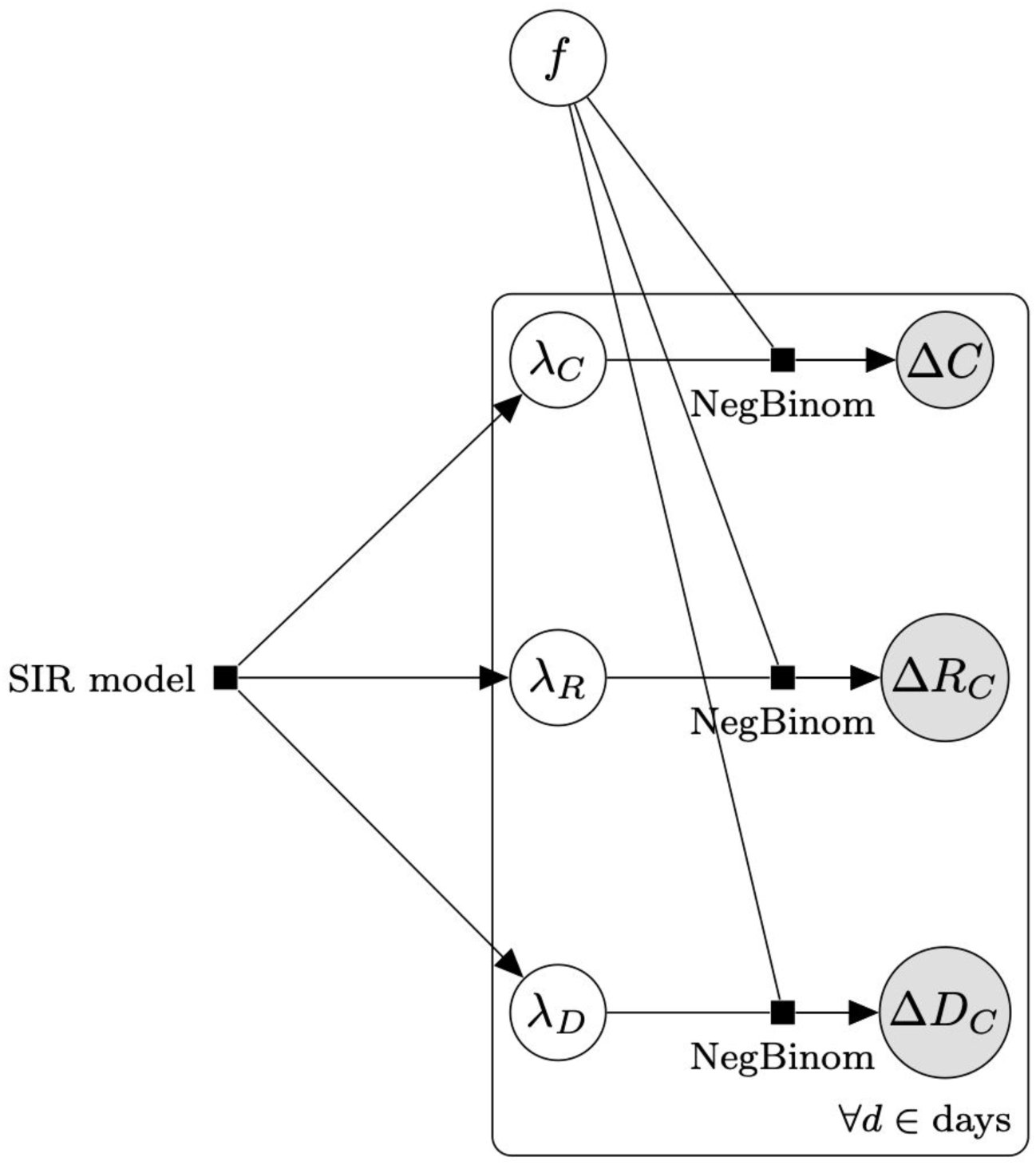
Diagram of generative observation model. The SIR models consist of differential equations that describe the evolution of the S, I, C, R_C_, and D_C_ variables. The models generate the time dependent rate of appearance of cases (*λ_C_*), case recoveries (*λ_R_*), and case deaths (*λ_D_*). These rates predict the observable data of daily new cases Δ*C*, new case recoveries Δ*R_C_*, and new case deaths Δ*D_C_* with a Negative Binomial likelihood function with an excess variance (*fλ*)^2^, compared to a Poisson likelihood. An expanded version of this diagram with additional details is available in Fig. S1.

Computer code written for this project is available at https://github.com/nih-niddk-mbs/covid-sicr.

## Results

Global estimates for R_t_, CAR_t_, and IFR_t_, for 25 representative countries (selected based on case counts) and US states with the prerequisite data, are shown in Figures 2–4 (see Supplementary Material Table S1 for every region analyzed). Overall, we found a consensus between the different model predictions, including between linear and nonlinear models (Supplementary Material Figure S2). Results presented are Bayesian LOO weighted model averages. CAR_t_ can be greater than 1 in rare cases where more cases were introduced by migration at outbreak onset than disappeared from the infected pool without being detected. Credible intervals were broad for most region specific estimates (see Supplementary Material Table S1).

Global average quantiles across all regions and models are shown in Figures 2–5. The models capture the dynamics of R_t_ (Fig. 2), CAR_t_ (Fig. 3), and IFR_t_ (Fig. 4). The global median R_0_ across all regions was 3.3 (C.I (1.5,8.3), Fig. 5). In the first week of the onset of COVID-19 appearance in a region (which differs by date for each region), we found that for every 1 case identified there are approximately 13 other infections unreported (CAR_t_ (week 0) of 0.07 (C.I (0.02,0.4)). The infection fatality ratio during the first week from regional onset has a global median of 0.0017 (C.I. (0.0005,0.009)) or 0.17%. As the infection spreads these estimates evolve over time such that the R_t_ on the week of April 15th, 2020 was 1.2 (C.I (0.6,2.5)). While R_t_ fell, the global median CAR_t_ and IFR_t_ increased over the same time period to 1 in 10 for CAR_t_ (April 15th, 2020) = 0.1 (C.I. (0.02,0.5)) (a larger fraction of infections ultimately detected) and 0.008 (C.I. (0.002,0.04)) or 0.8% for IFR_t_ (April 15th, 2020). IFR_t_ will be lower than the true mortality rate since it does not account for unreported deaths due to COVID-19. We have also not corrected for the fact that some of the current cases are critically ill and may die in the future. We should also note that since the onset date of COVID-19 varies regionally, the global median as of April 15 is an aggregate over different stages of regional pandemic progression.

We also evaluated the effects of quarantine and excess variability. Across models we observed a substantial decrease in infection rate by active cases. However, the residual infection rate was still above zero with broad uncertainty. The infectiousness of active cases (the known infected) across all regions was much lower than that of the remaining (unreported) infected (Figure 6), with a median global q = 0.03 (C.I. (0.001,0.16)). While the median global quarantine attenuation was large, there was a broad distribution with some areas having smaller degrees than others. Within Europe, Italy had the smallest median case infection rate consistent with strong quarantine measures (Pedersen and Meneghini) (see Supplementary Figure S3 for regional details).

Variability in reported data likely leads to overdispersion, i.e. to counts that are more variable than would be expected from a Poisson distribution. We thus accounted for and analyzed overdispersion by using a negative binomial distribution in which the excess variance is modeled as (*fλ*)^2^ where *λ* is the mean rate for that day and *f* is an overdispersion factor that we estimate. As shown in Supplementary Material Figure S4, *f* was approximately 1.5 across all regions.

**Figure 2:**
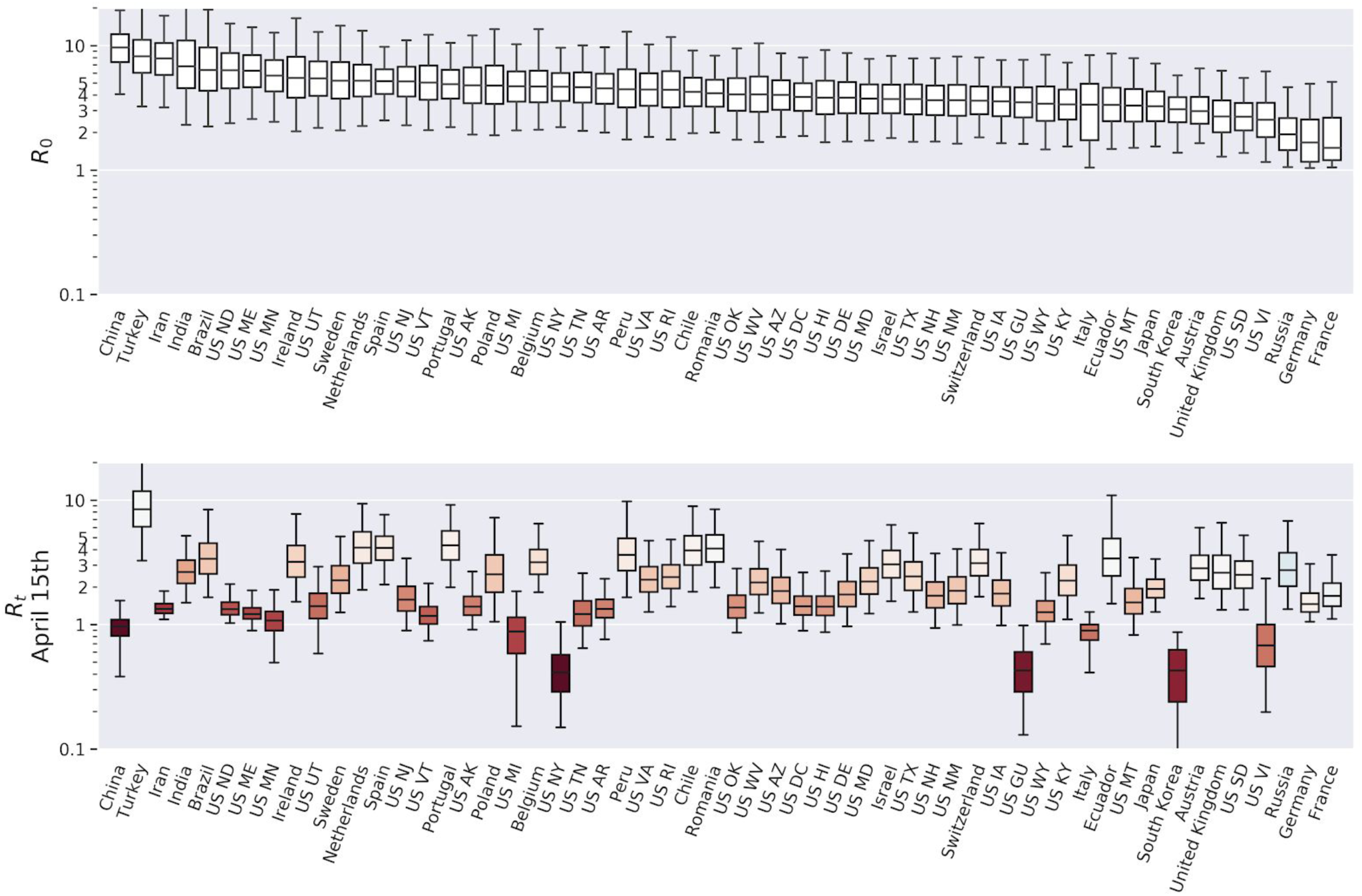
Estimates of the reproduction number *R_t_* across selected regions and time. Upper panel: *R_0_* for selected regions. Bars show median, interquartile range, and 95% credible interval for each region. Lower panel: R_t_ on April 15th in all regions. Color indicates change from R_0_ (more red is more reduction).

**Figure 3:**
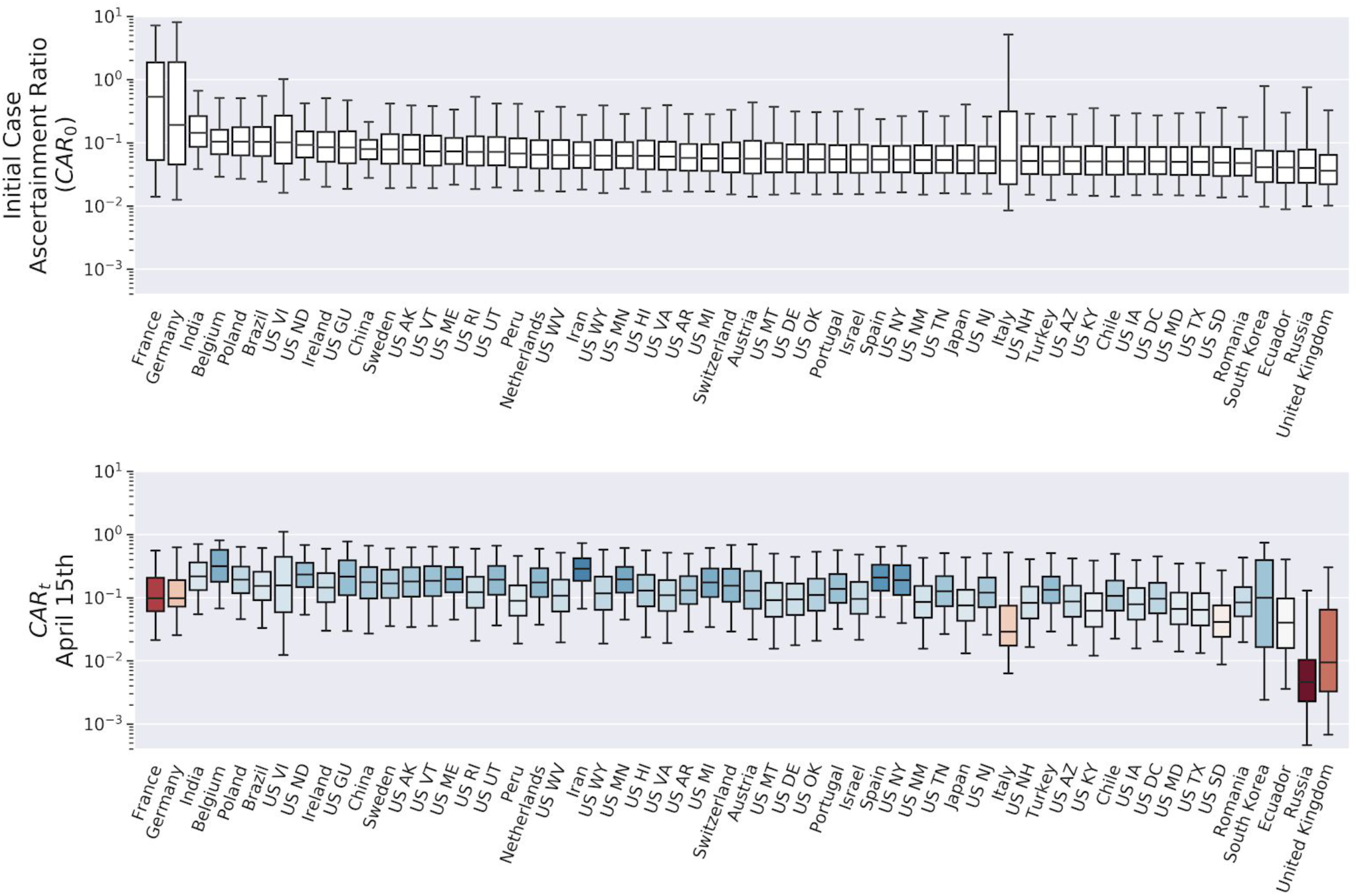
Estimates of the case ascertainment ratio (CAR_t_) across selected regions and time. Upper panel: Initial CAR_t_ for selected regions. Bars show median, interquartile range, and 95% credible interval for each region. Lower panel: CAR_t_ on April 15th in all regions. Color indicates change from initial CAR (more red is more reduction).

**Figure 4:**
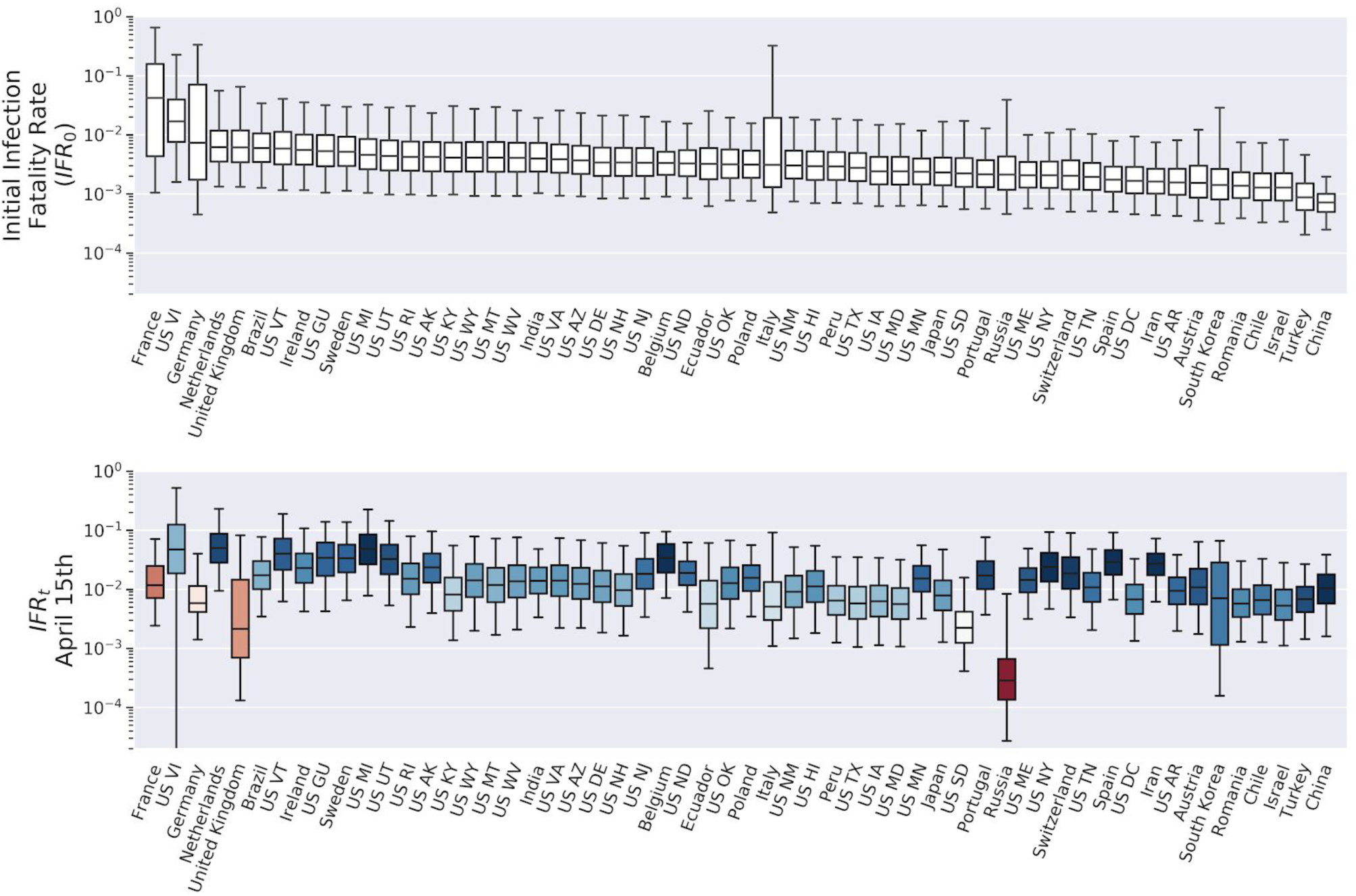
Estimates of the infection fatality rate IFR_t_ across selected regions and time. Upper panel: Initial IFR_t_ for selected regions. Bars show median, interquartile range, and 95% credible interval for each region. Lower panel: IFR_t_ on April 15th in all regions. Color indicates change from initial IFR (more red is more reduction).

**Figure 5:**
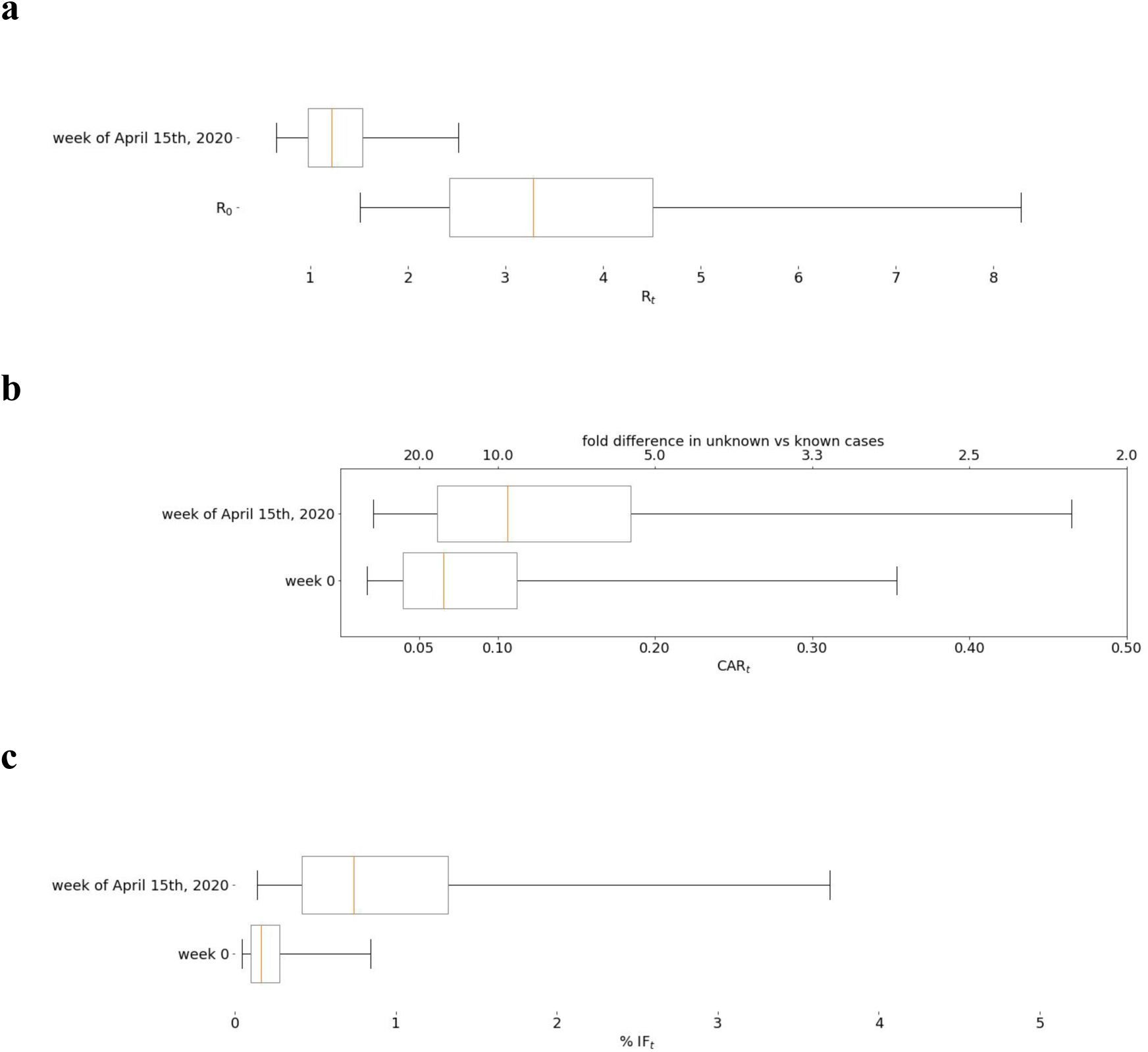
Global distributions of (a) R_t_, (b) CAR_t_, and (c) IFR_t_ using average across models. Figure shows quantiles median, interquartile range, and 95% credible interval.

**Figure 6:**
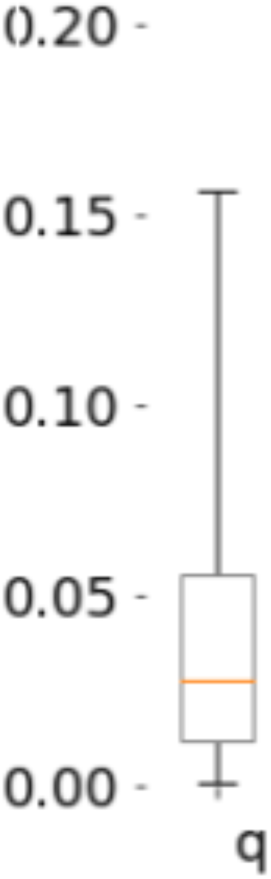
Effectiveness of quarantine. Shows a median q of 0.03 (C.I. (0.001, 0.16)) in the relative infection rate from active known cases. A value of q=0 would correspond to perfect quarantine of active eases (no new infections result from a case once it is identified). See Figure S2 for regional details.

## Discussion

We use our Bayesian mechanistic models to estimate the extent of COVID-19 prevalence, from publicly-available data. Importantly, we quantify the uncertainty of our estimates, finding them to be large in some regions. Our models perform this estimation task under simplified but plausible mechanisms for infection transmission, case detection, recovery, death, and disease spread mitigation.

The advantage of mechanistic models over non-mechanistic curve extrapolation (Murray and IHME COVID-19 health service utilization forecasting team) is that the model parameters have direct mechanistic interpretations that can be validated independently. More mechanisms and details can also be added incrementally.

### Estimate of the Case Ascertainment Ratio (CAR)

We show that the unaccounted cases exceed the reported by 10 to 20 fold, which is consistent with two independent groups in New York (one using PCR testing) (Sutton et al.; New York Times) but not with a similar test in Santa Clara (Bendavid et al.). However, factors such as false positive rates of the serological tests and selection bias could affect outcomes. Furthermore, the studies cited were not drawn from random samples of the population. Until wide-spread serological testing is implemented, the true proportion of cumulative infections will remain unknown, but we believe that our results provide a credible range for this proportion.

Our model could provide guidance on the effectiveness of various policies for reducing infectiousness until more complete data is obtained. Similar attempts have been made to model these factors. However, they have relied on priors for R_0_ based on confirmed cases (Lourenco et al.; Flaxman et al.; Ferguson et al.), restricted data to reported deaths or cases only (Flaxman et al.; Pedersen and Meneghini; Murray and IHME COVID-19 health service utilization forecasting team), made strong assumptions about quarantine (Pedersen and Meneghini), or ignored the underlying mechanisms that guide the dynamics of epidemics (Murray and IHME COVID-19 health service utilization forecasting team). We are able to improve identifiability of key parameters compared to previous results as in (Lourenco et al.) because we use more information. The motivation to use only case death data is understandable as it may be a more reliable measure than case counts but it is insufficient to constrain the dynamics alone and thus has much less predictive power. However, as shown in the Supplementary Material (Sec. S2), models that includie cases, case recoveries, and case deaths, provide sufficient information for identifiability of undocumented infections although the uncertainty can be large.

### Estimate of the time-dependent reproduction number (R_t_)

Our model predicts the time dependent reproduction number R_t_ for each region, which can give an estimate as to how well mitigation policies are working. Our results are consistent with social distancing, or other policies reducing transmission, as shown by the decrease in R_t_ relative to R_0_ at outbreak onset. Our directly inferred region-wide R_0_ estimate of 3.3 is larger than the mean of priors chosen for some other models (Lourenco et al.; Flaxman et al.; Ferguson et al.), but it is consistent with estimates from other work (Cao et al.; Sanche et al.) including several reviewed in (Liu et al.). Our estimate is close to the upper estimates for Italy by Pedersen and Menegheni (Pedersen and Meneghini) of 3.5 when removing quarantine on active cases. Using a similar linear SIR model with complete quarantine lowered their R_0_ estimate to 2.6. Our estimate of R_0_ was robust to assumptions on the effectiveness of quarantine (models without this factor gave similar estimates for R_0_ see Supplementary Material Figure S2). Our estimate of parameter q near zero for Italy implies near perfect quarantine. There was more incomplete quarantine in other regions.

### Policy implications

We found a decrease in R_t_ over time with the global estimate currently near 1, though many regions are still well above this as of April 15. However, R_t_ does not need to be below 1 for herd immunity to stop the spread of COVID-19. When the fraction of infected individuals reaches 1-1/R_t_ regionally, then in the absence of reinfection from outside, herd immunity will quench the pandemic locally (Fine et al.). Although there are regional variations, if the current estimate of R_t_ persists then the pandemic will begin to extinguish itself when 17% (C.I. (0%, 60%)) of the susceptible global population has achieved resistance due to either recovery from prior infection or vaccination (0% means we have herd immunity now, under the current mitigation measures). However, if mitigation measures are lifted and R_t_ returns to R_0_, then that fraction rises to 70% (C.I. (33%, 88%)) to guard against further waves of illness upon reinfection.

The time-varying models give other notable results. On average CAR_t_ has increased, indicating that a larger proportion of infections are being detected. This might be explained by recognition of symptoms increasing during the pandemic coupled with broader availability of testing. Our estimate of IFR_t_ also increased during the spread of the infection. This is probably due to the fact that early in the pandemic those fated to die would have been less likely to be identified as COVID-19 deaths, or more generally from changes in how deaths are reported over time. Because the IFR that we report uses the conventional definition, it is “right censored” -- at any given time point the number of deaths from the current number of infections will continue to grow for some time -- it could reflect an underestimate of the true mortality rate. Because our models explicitly estimate the rate at which both case and non-case infected either die or recover, one can derive a closed form expression for the true mortality rate.

### Assumptions and Limitations

Driven by mechanistic considerations, we make several simplifying modeling assumptions that we believe to be reasonable. Chiefly, we make the standard assumptions of SIR models. For instance, we ignore heterogeneity within populations and the effect of connectivity between populations. We subsume heterogeneity of populations themselves into the model parameters, which are to be interpreted as population-averaged quantities. While mitigation measures have greatly reduced travel, cross-population transmission may still be occurring at low levels. Although our models account for mitigation and for diagnostic changes, our rigid implementation of these factors is unlikely to fully capture the true time course of their effects. Finally, biologically, our model considers an extremely simplified version of disease progression within individuals, ignoring time inhomogeneous mechanisms that may be significant (Bottcher et al.).

Motivated by statistical considerations, we also consider reduced linear approximations to the SIR models, where changes in the size of the susceptible pool are ignored. Empirically, we find these approximations to be more stable for fitting the pandemic in the early stages than the standard nonlinear SIR model. A complication in using the standard nonlinear SIR model lies in the knowledge of the size of the susceptible population. In addition to the fact that it is not known what fraction of the population has innate immunity, the inherent heterogeneity of the spread of disease in the early stages renders the susceptible population estimate problematic if not impossible. (In our nonlinear models we fit it as an adjustable parameter). The linearized model circumvents this difficulty by scaling out the population, and to some degree ameliorates the homogeneous assumption, by averaging over the initial growth of the local clusters. Hence, for the linearized model during the early stages of an epidemic, heterogeneity in infectiousness has less of an impact on the underlying aggregate state variables, and so does not provide a worse fit than a well-mixed population with the same observed data. Only later in the epidemic when the effects of herd immunity are important will the limitations of a linear model with homogeneous mixing be revealed, although the time dependence in the infectiousness we introduce could also be capturing the effects of herd immunity.

Outside of our modeling assumptions, issues of data quality may affect the accuracy of our estimates. Disparate criteria exist across regions for defining cases and case deaths, oftentimes changing within regions themselves as each epidemic progresses.

Additionally, data (including death counts) reported by many regions is likely incomplete or unreliable, as evidenced in large spikes of unexplained deaths not officially recognized as Covid-19 deaths *(Covid-19 Data - Tracking Covid-19 Excess Deaths across Countries* | *Graphic Detail* | *The Economist)*; while we believe that systematic under-testing can be overcome up to a point, totally inconsistent or fraudulent reporting cannot be.

### Future Extensions

Additional factors need to be explored in future models. For example, a more robust and flexible implementation of mitigation, as well as heterogeneity of disease progression within individuals and in their social interactions, is essential. Rather than fitting regions independently it should be possible to fit a hierarchical global model in which region-specific parameters are partially pooled to achieve additional regularization and to narrow credible intervals. The linear model is robust to uncertainty in the size of the initial susceptible population but if the disease progresses such that we begin to reach population saturation, we must use a nonlinear model that explicitly accounts for population size to make reliable parameter estimates although the time dependence in infectiousness can partially mimic this nonlinear effect. We are optimistic that such a transition will be seamless since we find consistency in model predictions between linear and nonlinear models in the current work.

Definitions

Definitions
| Term | Meaning |
| --- | --- |
| $R_t$ | Reproduction number at time $t$: the average number of secondary infected individuals due to a single infected individual |
| $CFR_t$ | Case ascertainment ratio at time $t$: the cumulative number of cases reported over cumulative number of infections (observed or not) |
| $IFR_t$ | Infection fatality ratio at time $t$: the cumulative number of known deaths over the cumulative number of infections (observed or not)<br>*This will be lower than the mortality rate, as mortality includes unobserved deaths due to infection.* |
| $q$ | Relative infection rate by known active cases: this is relative to the baseline infection rate of unknown infected cases (including asymptomatic carriers). A value of $q = 0$ is perfect quarantine of active cases. |
| Over-Dispersion Factor, f | Excess variability in counts beyond a Poisson process, modeled as $\text{variance} = \text{mean} + (f * \text{mean})^2$ |
| Credible interval | An interval spanning the middle 95% of the (posterior) distribution for the estimates, i.e. 2.5th to 97.5th percentile |

## Data Availability

All data and computer code used is publicly available

https://github.com/nih-niddk-mbs/covid-sicr

## Acknowledgments

The authors would like to thank Christopher Kim for early assistance and Kevin Hall for critical comments. CCC and SV were supported by the Intramural Program of the NIH, NIDDK. RCG was supported by NIDCD, NINDS, and NSF. This work utilized the computational resources of the NIH HPC Biowulf cluster (http://hpc.nih.gov).

## Supplementary Material

**Table S1.**
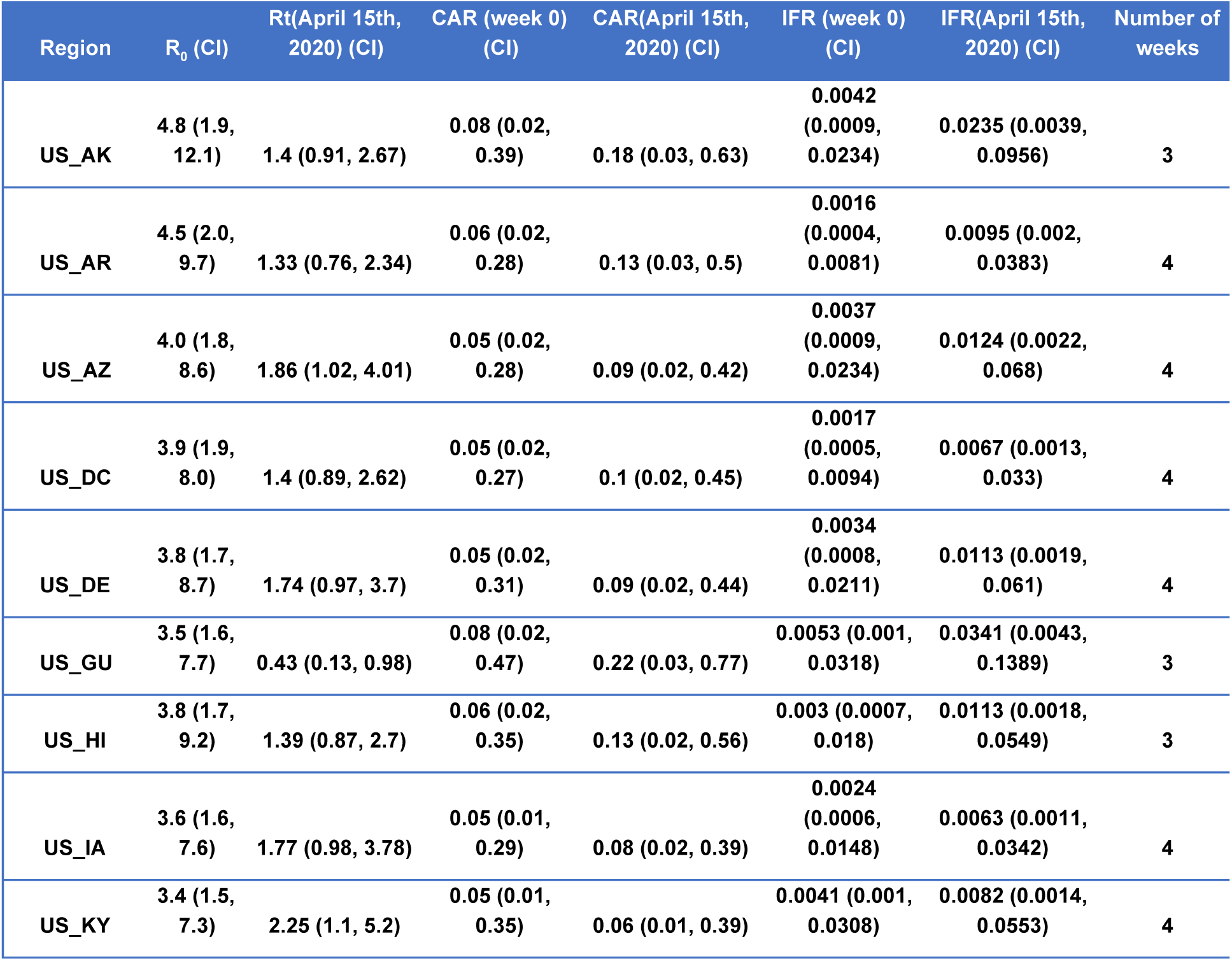
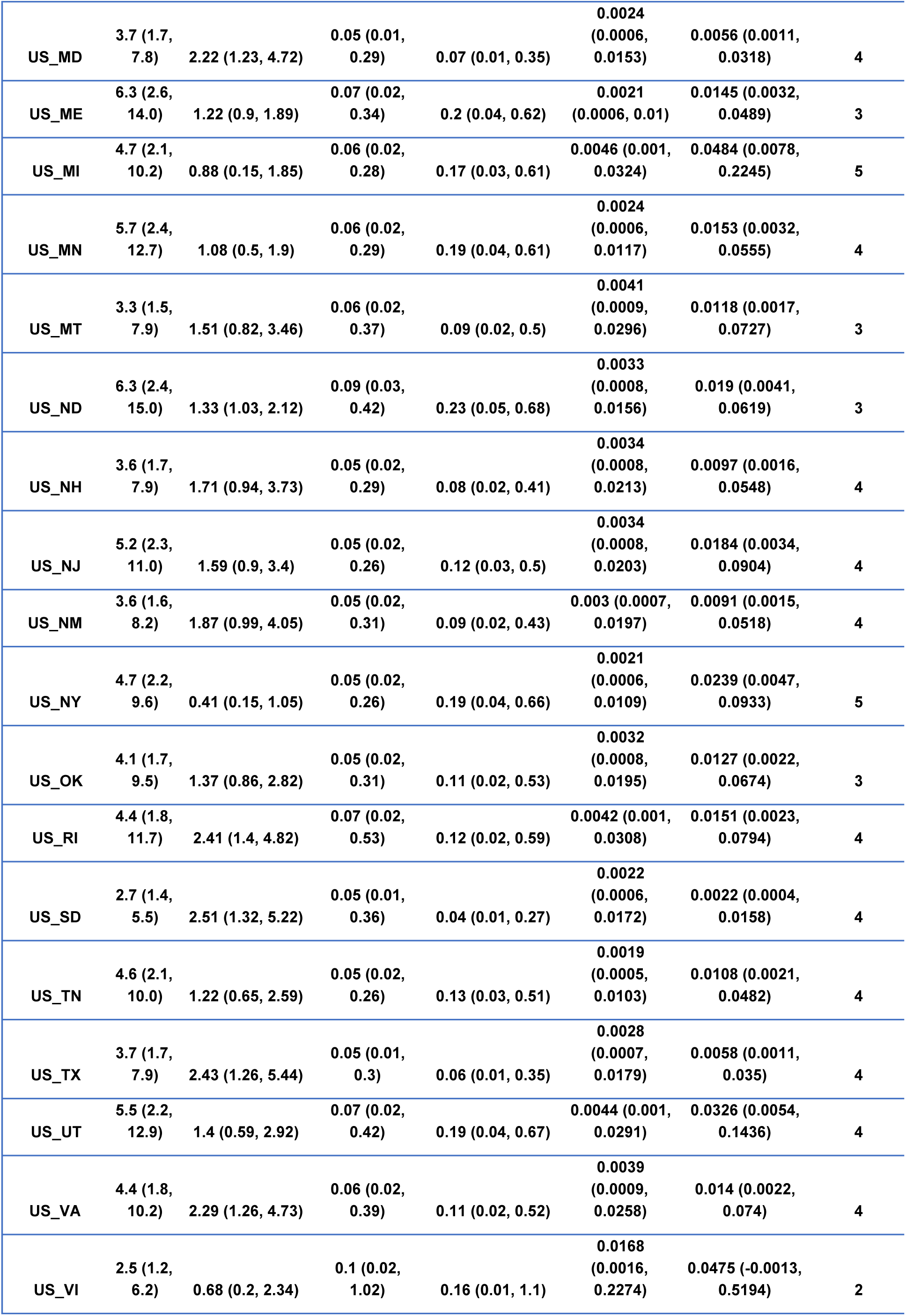
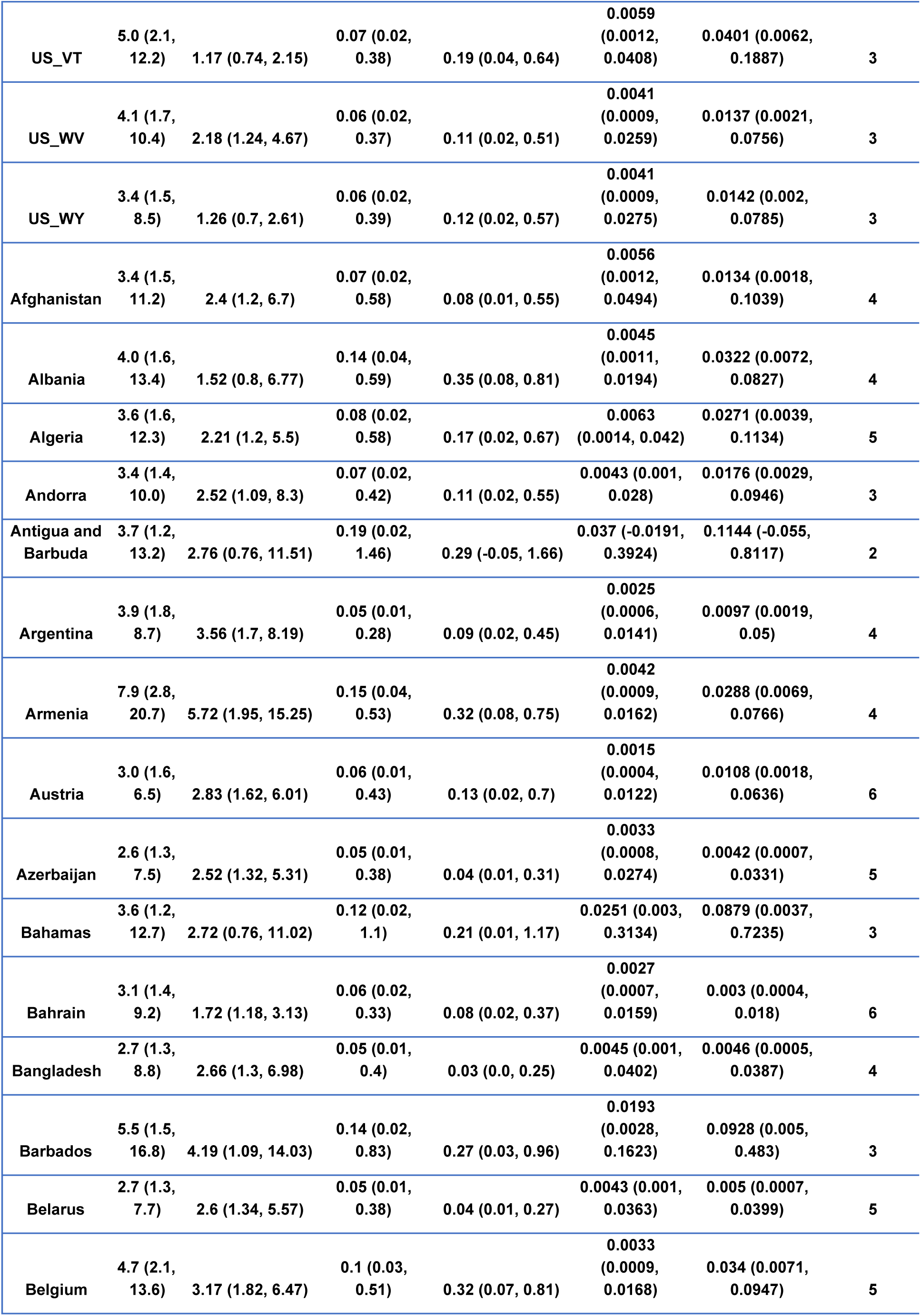
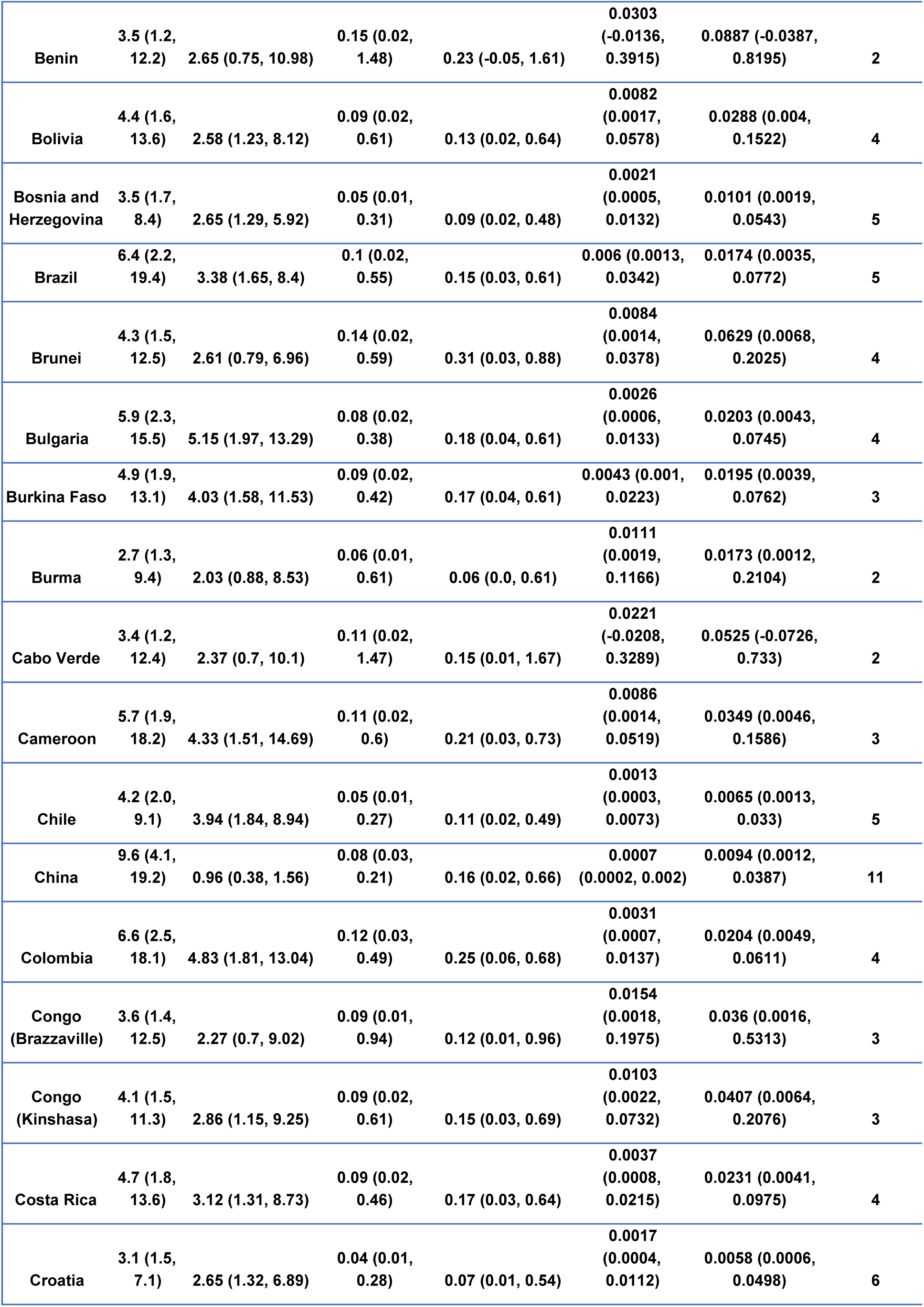
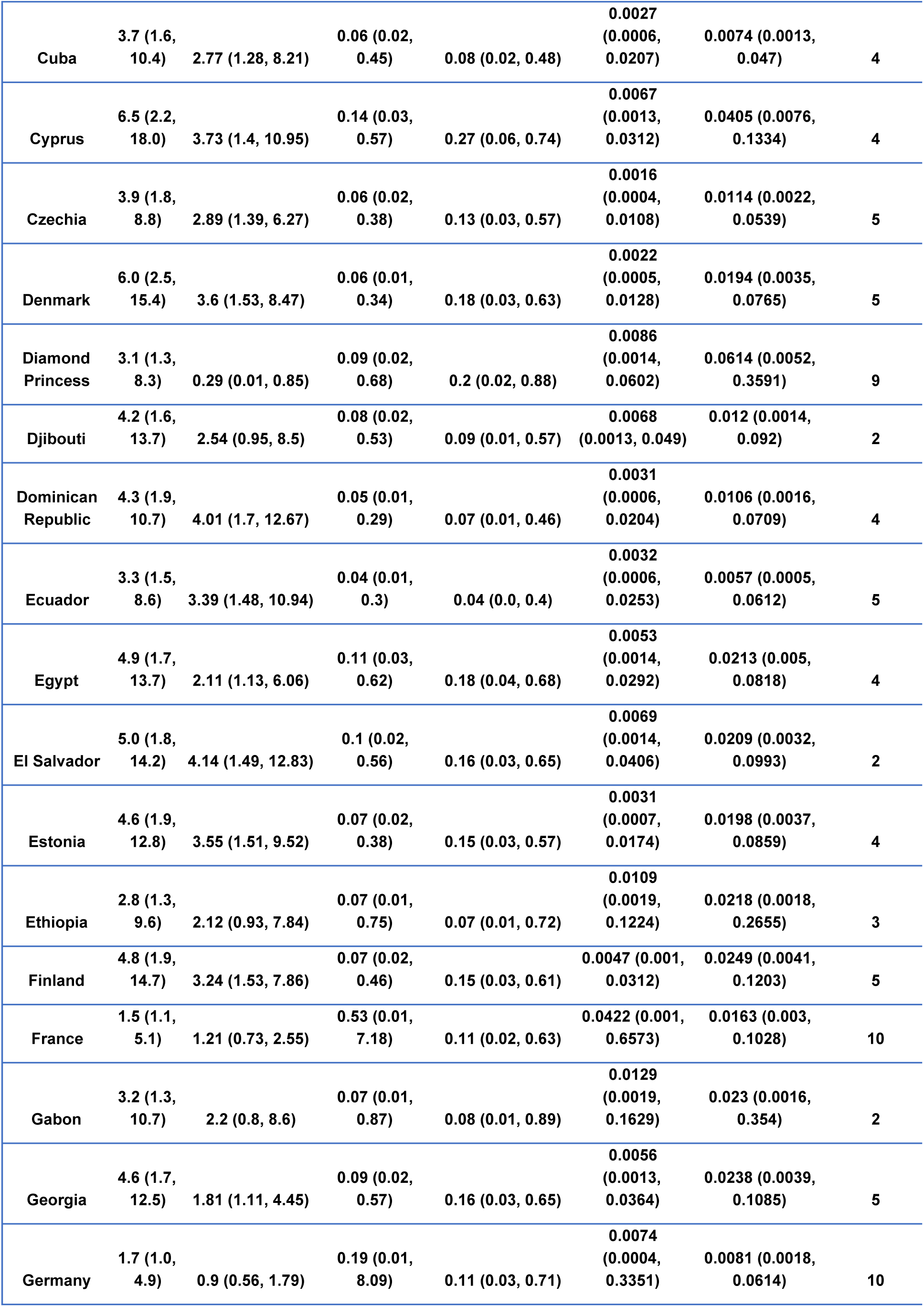
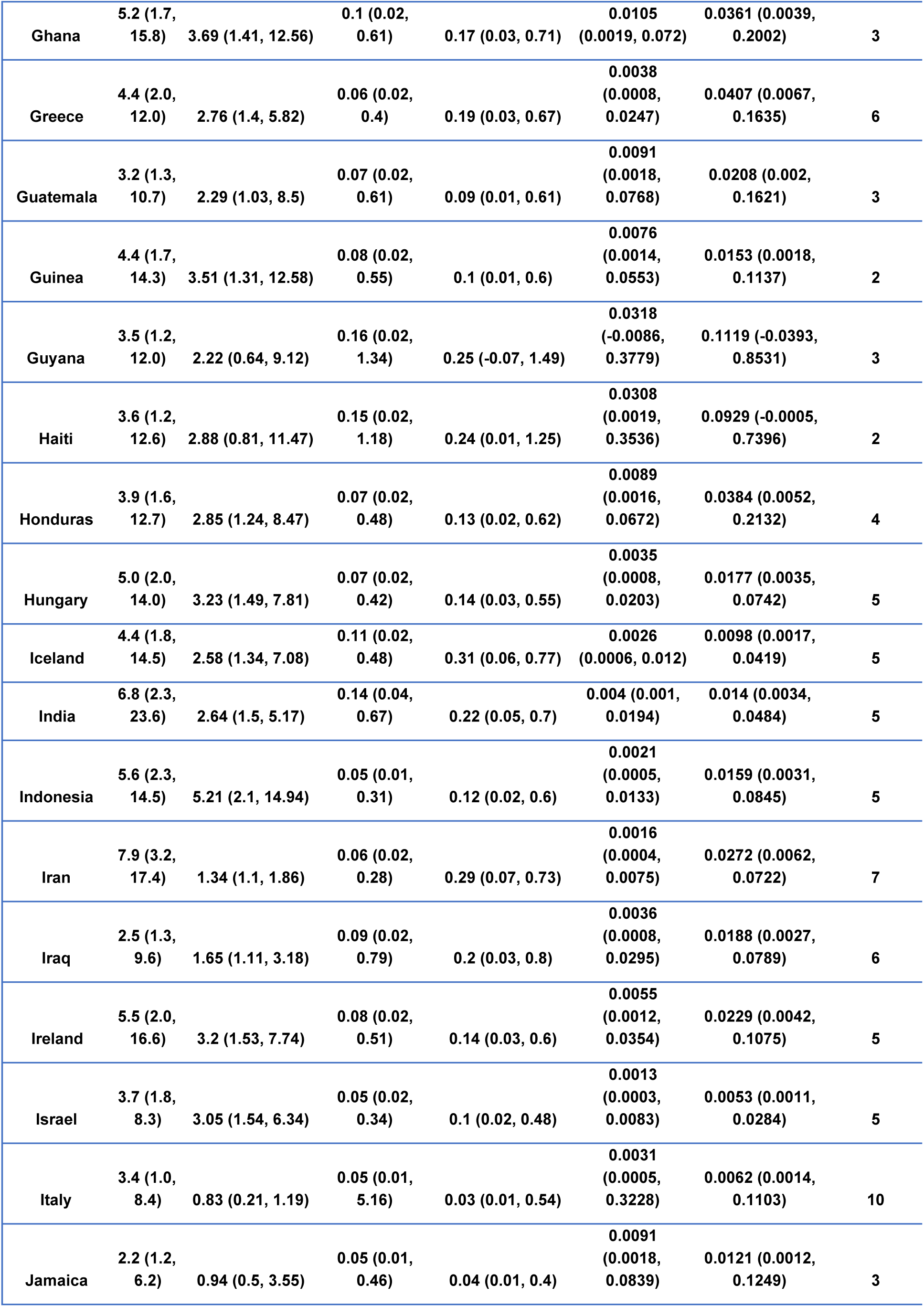
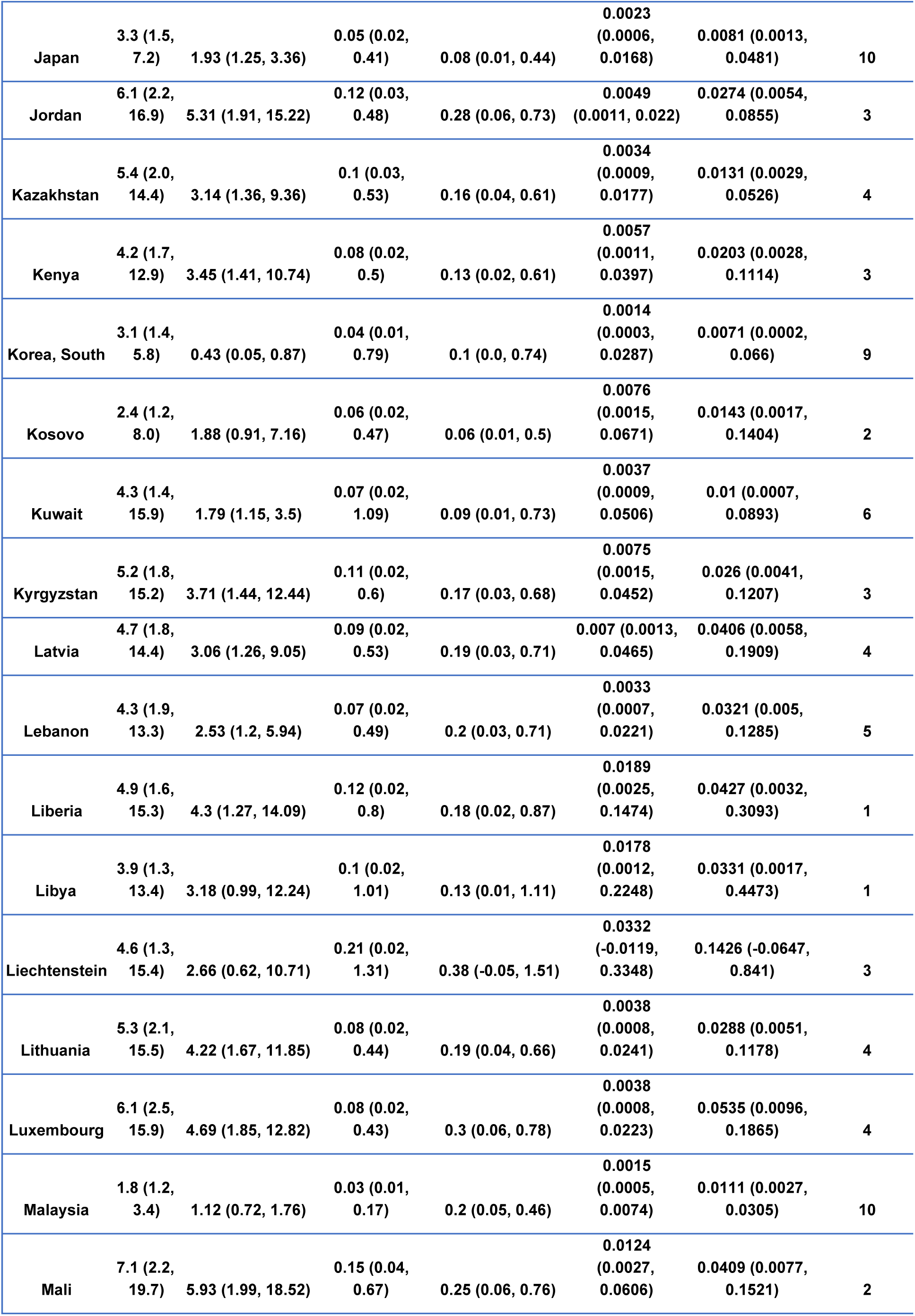
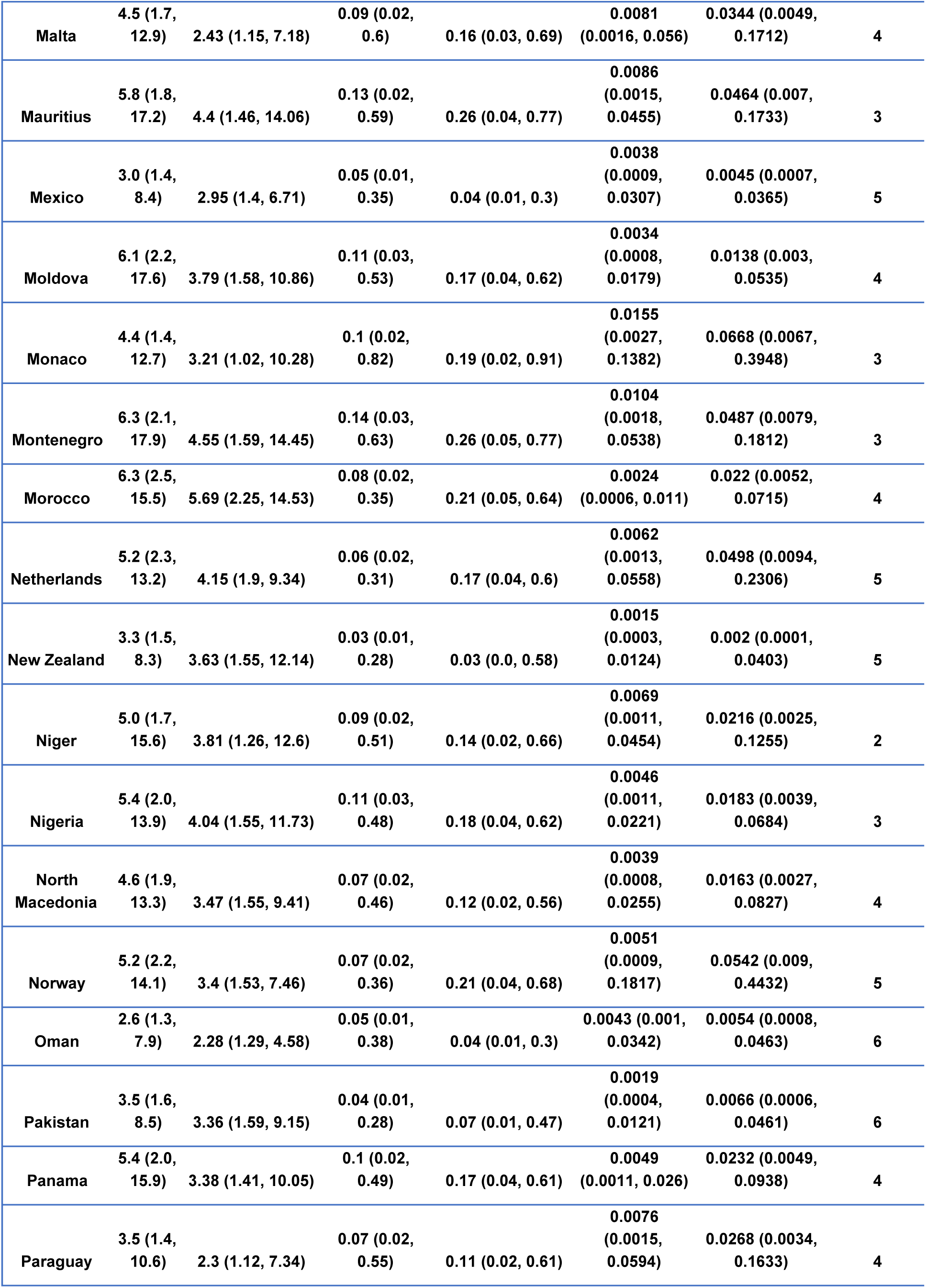
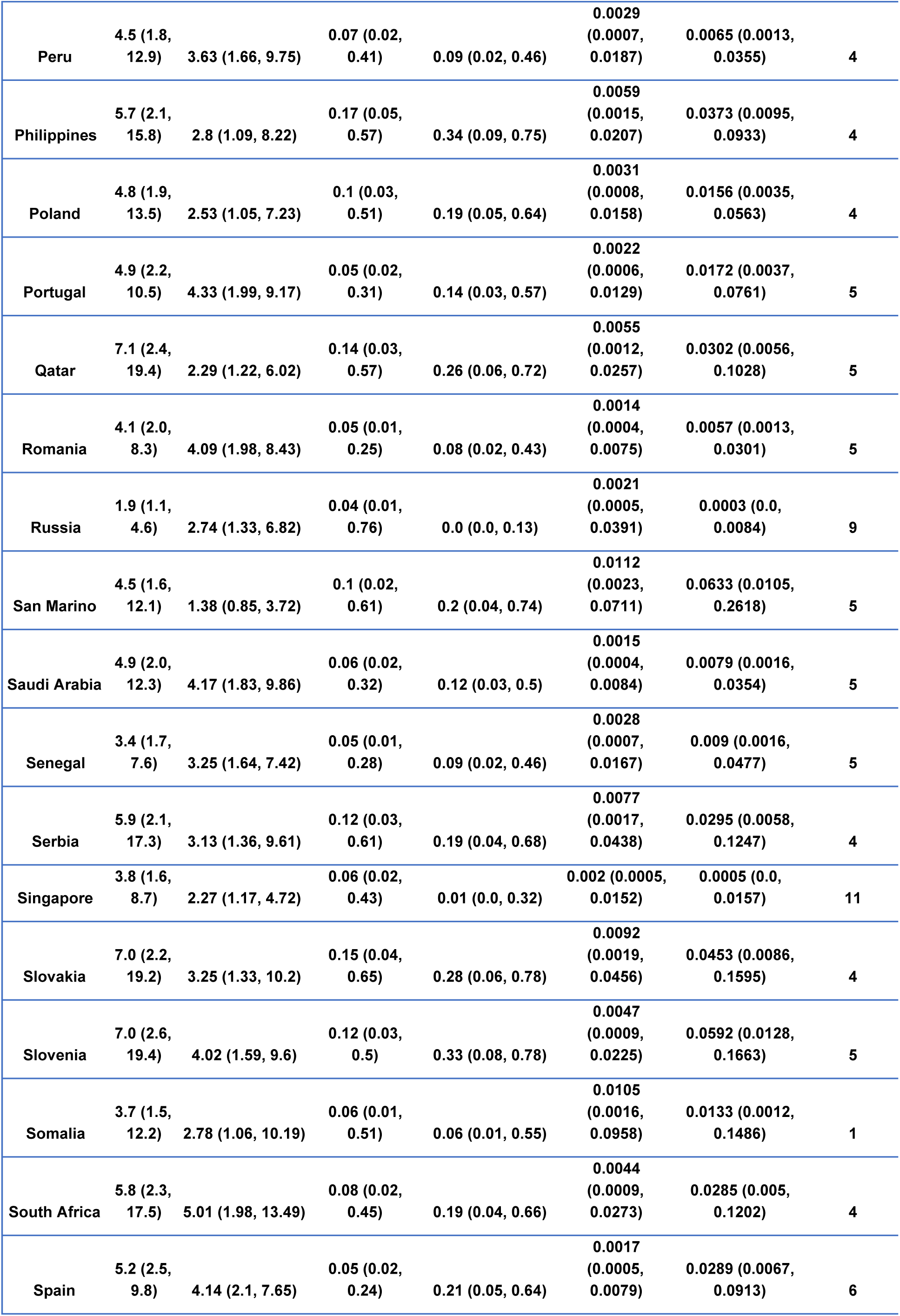
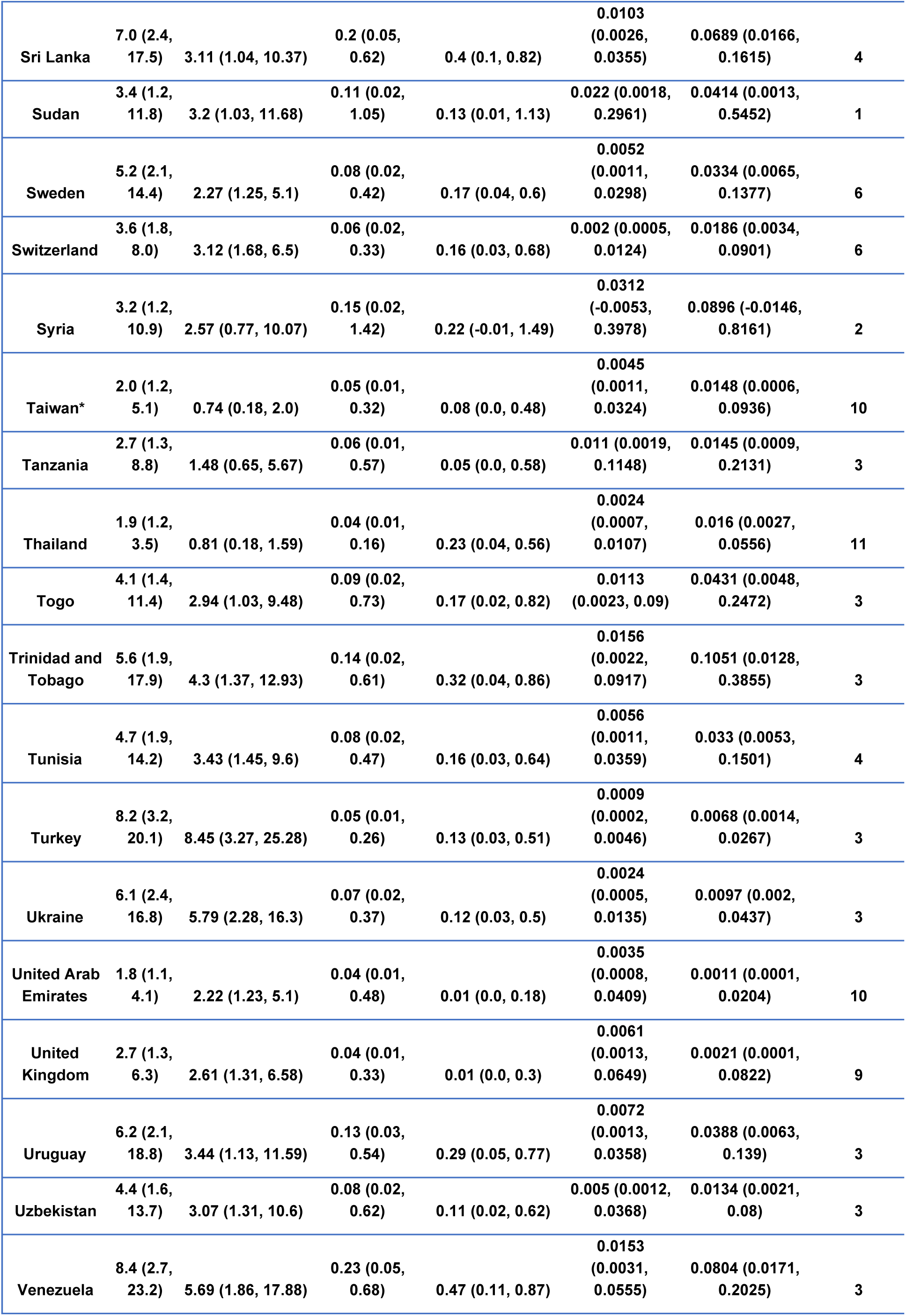
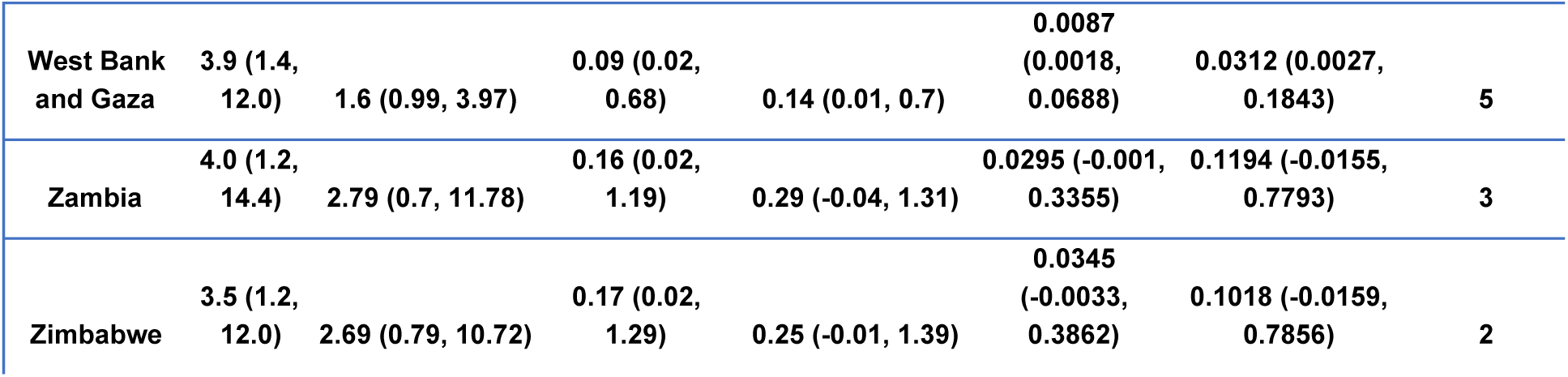

| Region | $R_0$ (CI) | Rt(April 15th, 2020) (CI) | CAR (week 0) (CI) | CAR(April 15th, 2020) (CI) | IFR (week 0) (CI) | IFR(April 15th, 2020) (CI) | Number of weeks |
| --- | --- | --- | --- | --- | --- | --- | --- |
| US_AK | 4.8 (1.9, 12.1) | 1.4 (0.91, 2.67) | 0.08 (0.02, 0.39) | 0.18 (0.03, 0.63) | 0.0042 (0.0009, 0.0234) | 0.0235 (0.0039, 0.0956) | 3 |
| US_AR | 4.5 (2.0, 9.7) | 1.33 (0.76, 2.34) | 0.06 (0.02, 0.28) | 0.13 (0.03, 0.5) | 0.0016 (0.0004, 0.0081) | 0.0095 (0.002, 0.0383) | 4 |
| US_AZ | 4.0 (1.8, 8.6) | 1.86 (1.02, 4.01) | 0.05 (0.02, 0.28) | 0.09 (0.02, 0.42) | 0.0037 (0.0009, 0.0234) | 0.0124 (0.0022, 0.068) | 4 |
| US_DC | 3.9 (1.9, 8.0) | 1.4 (0.89, 2.62) | 0.05 (0.02, 0.27) | 0.1 (0.02, 0.45) | 0.0017 (0.0005, 0.0094) | 0.0067 (0.0013, 0.033) | 4 |
| US_DE | 3.8 (1.7, 8.7) | 1.74 (0.97, 3.7) | 0.05 (0.02, 0.31) | 0.09 (0.02, 0.44) | 0.0034 (0.0008, 0.0211) | 0.0113 (0.0019, 0.061) | 4 |
| US_GU | 3.5 (1.6, 7.7) | 0.43 (0.13, 0.98) | 0.08 (0.02, 0.47) | 0.22 (0.03, 0.77) | 0.0053 (0.001, 0.0318) | 0.0341 (0.0043, 0.1389) | 3 |
| US_HI | 3.8 (1.7, 9.2) | 1.39 (0.87, 2.7) | 0.06 (0.02, 0.35) | 0.13 (0.02, 0.56) | 0.003 (0.0007, 0.018) | 0.0113 (0.0018, 0.0549) | 3 |
| US_IA | 3.6 (1.6, 7.6) | 1.77 (0.98, 3.78) | 0.05 (0.01, 0.29) | 0.08 (0.02, 0.39) | 0.0024 (0.0006, 0.0148) | 0.0063 (0.0011, 0.0342) | 4 |
| US_KY | 3.4 (1.5, 7.3) | 2.25 (1.1, 5.2) | 0.05 (0.01, 0.35) | 0.06 (0.01, 0.39) | 0.0041 (0.001, 0.0308) | 0.0082 (0.0014, 0.0553) | 4 |
| US_MD | 3.7 (1.7, 7.8) | 2.22 (1.23, 4.72) | 0.05 (0.01, 0.29) | 0.07 (0.01, 0.35) | 0.0024 (0.0006, 0.0153) | 0.0056 (0.0011, 0.0318) | 4 |
| US_ME | 6.3 (2.6, 14.0) | 1.22 (0.9, 1.89) | 0.07 (0.02, 0.34) | 0.2 (0.04, 0.62) | 0.0021 (0.0006, 0.01) | 0.0145 (0.0032, 0.0489) | 3 |
| US_MI | 4.7 (2.1, 10.2) | 0.88 (0.15, 1.85) | 0.06 (0.02, 0.28) | 0.17 (0.03, 0.61) | 0.0046 (0.001, 0.0324) | 0.0484 (0.0078, 0.2245) | 5 |
| US_MN | 5.7 (2.4, 12.7) | 1.08 (0.5, 1.9) | 0.06 (0.02, 0.29) | 0.19 (0.04, 0.61) | 0.0024 (0.0006, 0.0117) | 0.0153 (0.0032, 0.0555) | 4 |
| US_MT | 3.3 (1.5, 7.9) | 1.51 (0.82, 3.46) | 0.06 (0.02, 0.37) | 0.09 (0.02, 0.5) | 0.0041 (0.0009, 0.0296) | 0.0118 (0.0017, 0.0727) | 3 |
| US_ND | 6.3 (2.4, 15.0) | 1.33 (1.03, 2.12) | 0.09 (0.03, 0.42) | 0.23 (0.05, 0.68) | 0.0033 (0.0008, 0.0156) | 0.019 (0.0041, 0.0619) | 3 |
| US_NH | 3.6 (1.7, 7.9) | 1.71 (0.94, 3.73) | 0.05 (0.02, 0.29) | 0.08 (0.02, 0.41) | 0.0034 (0.0008, 0.0213) | 0.0097 (0.0016, 0.0548) | 4 |
| US_NJ | 5.2 (2.3, 11.0) | 1.59 (0.9, 3.4) | 0.05 (0.02, 0.26) | 0.12 (0.03, 0.5) | 0.0034 (0.0008, 0.0203) | 0.0184 (0.0034, 0.0904) | 4 |
| US_NM | 3.6 (1.6, 8.2) | 1.87 (0.99, 4.05) | 0.05 (0.02, 0.31) | 0.09 (0.02, 0.43) | 0.003 (0.0007, 0.0197) | 0.0091 (0.0015, 0.0518) | 4 |
| US_NY | 4.7 (2.2, 9.6) | 0.41 (0.15, 1.05) | 0.05 (0.02, 0.26) | 0.19 (0.04, 0.66) | 0.0021 (0.0006, 0.0109) | 0.0239 (0.0047, 0.0933) | 5 |
| US_OK | 4.1 (1.7, 9.5) | 1.37 (0.86, 2.82) | 0.05 (0.02, 0.31) | 0.11 (0.02, 0.53) | 0.0032 (0.0008, 0.0195) | 0.0127 (0.0022, 0.0674) | 3 |
| US_RI | 4.4 (1.8, 11.7) | 2.41 (1.4, 4.82) | 0.07 (0.02, 0.53) | 0.12 (0.02, 0.59) | 0.0042 (0.001, 0.0308) | 0.0151 (0.0023, 0.0794) | 4 |
| US_SD | 2.7 (1.4, 5.5) | 2.51 (1.32, 5.22) | 0.05 (0.01, 0.36) | 0.04 (0.01, 0.27) | 0.0022 (0.0006, 0.0172) | 0.0022 (0.0004, 0.0158) | 4 |
| US_TN | 4.6 (2.1, 10.0) | 1.22 (0.65, 2.59) | 0.05 (0.02, 0.26) | 0.13 (0.03, 0.51) | 0.0019 (0.0005, 0.0103) | 0.0108 (0.0021, 0.0482) | 4 |
| US_TX | 3.7 (1.7, 7.9) | 2.43 (1.26, 5.44) | 0.05 (0.01, 0.3) | 0.06 (0.01, 0.35) | 0.0028 (0.0007, 0.0179) | 0.0058 (0.0011, 0.035) | 4 |
| US_UT | 5.5 (2.2, 12.9) | 1.4 (0.59, 2.92) | 0.07 (0.02, 0.42) | 0.19 (0.04, 0.67) | 0.0044 (0.001, 0.0291) | 0.0326 (0.0054, 0.1436) | 4 |
| US_VA | 4.4 (1.8, 10.2) | 2.29 (1.26, 4.73) | 0.06 (0.02, 0.39) | 0.11 (0.02, 0.52) | 0.0039 (0.0009, 0.0258) | 0.014 (0.0022, 0.074) | 4 |
| US_VI | 2.5 (1.2, 6.2) | 0.68 (0.2, 2.34) | 0.1 (0.02, 1.02) | 0.16 (0.01, 1.1) | 0.0168 (0.0016, 0.2274) | 0.0475 (-0.0013, 0.5194) | 2 |
| US_VT | 5.0 (2.1, 12.2) | 1.17 (0.74, 2.15) | 0.07 (0.02, 0.38) | 0.19 (0.04, 0.64) | 0.0059 (0.0012, 0.0408) | 0.0401 (0.0062, 0.1887) | 3 |
| US_WV | 4.1 (1.7, 10.4) | 2.18 (1.24, 4.67) | 0.06 (0.02, 0.37) | 0.11 (0.02, 0.51) | 0.0041 (0.0009, 0.0259) | 0.0137 (0.0021, 0.0756) | 3 |
| US_WY | 3.4 (1.5, 8.5) | 1.26 (0.7, 2.61) | 0.06 (0.02, 0.39) | 0.12 (0.02, 0.57) | 0.0041 (0.0009, 0.0275) | 0.0142 (0.002, 0.0785) | 3 |
| Afghanistan | 3.4 (1.5, 11.2) | 2.4 (1.2, 6.7) | 0.07 (0.02, 0.58) | 0.08 (0.01, 0.55) | 0.0056 (0.0012, 0.0494) | 0.0134 (0.0018, 0.1039) | 4 |
| Albania | 4.0 (1.6, 13.4) | 1.52 (0.8, 6.77) | 0.14 (0.04, 0.59) | 0.35 (0.08, 0.81) | 0.0045 (0.0011, 0.0194) | 0.0322 (0.0072, 0.0827) | 4 |
| Algeria | 3.6 (1.6, 12.3) | 2.21 (1.2, 5.5) | 0.08 (0.02, 0.58) | 0.17 (0.02, 0.67) | 0.0063 (0.0014, 0.042) | 0.0271 (0.0039, 0.1134) | 5 |
| Andorra | 3.4 (1.4, 10.0) | 2.52 (1.09, 8.3) | 0.07 (0.02, 0.42) | 0.11 (0.02, 0.55) | 0.0043 (0.001, 0.028) | 0.0176 (0.0029, 0.0946) | 3 |
| Antigua and Barbuda | 3.7 (1.2, 13.2) | 2.76 (0.76, 11.51) | 0.19 (0.02, 1.46) | 0.29 (-0.05, 1.66) | 0.037 (-0.0191, 0.3924) | 0.1144 (-0.055, 0.8117) | 2 |
| Argentina | 3.9 (1.8, 8.7) | 3.56 (1.7, 8.19) | 0.05 (0.01, 0.28) | 0.09 (0.02, 0.45) | 0.0025 (0.0006, 0.0141) | 0.0097 (0.0019, 0.05) | 4 |
| Armenia | 7.9 (2.8, 20.7) | 5.72 (1.95, 15.25) | 0.15 (0.04, 0.53) | 0.32 (0.08, 0.75) | 0.0042 (0.0009, 0.0162) | 0.0288 (0.0069, 0.0766) | 4 |
| Austria | 3.0 (1.6, 6.5) | 2.83 (1.62, 6.01) | 0.06 (0.01, 0.43) | 0.13 (0.02, 0.7) | 0.0015 (0.0004, 0.0122) | 0.0108 (0.0018, 0.0636) | 6 |
| Azerbaijan | 2.6 (1.3, 7.5) | 2.52 (1.32, 5.31) | 0.05 (0.01, 0.38) | 0.04 (0.01, 0.31) | 0.0033 (0.0008, 0.0274) | 0.0042 (0.0007, 0.0331) | 5 |
| Bahamas | 3.6 (1.2, 12.7) | 2.72 (0.76, 11.02) | 0.12 (0.02, 1.1) | 0.21 (0.01, 1.17) | 0.0251 (0.003, 0.3134) | 0.0879 (0.0037, 0.7235) | 3 |
| Bahrain | 3.1 (1.4, 9.2) | 1.72 (1.18, 3.13) | 0.06 (0.02, 0.33) | 0.08 (0.02, 0.37) | 0.0027 (0.0007, 0.0159) | 0.003 (0.0004, 0.018) | 6 |
| Bangladesh | 2.7 (1.3, 8.8) | 2.66 (1.3, 6.98) | 0.05 (0.01, 0.4) | 0.03 (0.0, 0.25) | 0.0045 (0.001, 0.0402) | 0.0046 (0.0005, 0.0387) | 4 |
| Barbados | 5.5 (1.5, 16.8) | 4.19 (1.09, 14.03) | 0.14 (0.02, 0.83) | 0.27 (0.03, 0.96) | 0.0193 (0.0028, 0.1623) | 0.0928 (0.005, 0.483) | 3 |
| Belarus | 2.7 (1.3, 7.7) | 2.6 (1.34, 5.57) | 0.05 (0.01, 0.38) | 0.04 (0.01, 0.27) | 0.0043 (0.001, 0.0363) | 0.005 (0.0007, 0.0399) | 5 |
| Belgium | 4.7 (2.1, 13.6) | 3.17 (1.82, 6.47) | 0.1 (0.03, 0.51) | 0.32 (0.07, 0.81) | 0.0033 (0.0009, 0.0168) | 0.034 (0.0071, 0.0947) | 5 |
| Benin | 3.5 (1.2, 12.2) | 2.65 (0.75, 10.98) | 0.15 (0.02, 1.48) | 0.23 (-0.05, 1.61) | 0.0303 (-0.0136, 0.3915) | 0.0887 (-0.0387, 0.8195) | 2 |
| Bolivia | 4.4 (1.6, 13.6) | 2.58 (1.23, 8.12) | 0.09 (0.02, 0.61) | 0.13 (0.02, 0.64) | 0.0082 (0.0017, 0.0578) | 0.0288 (0.004, 0.1522) | 4 |
| Bosnia and Herzegovina | 3.5 (1.7, 8.4) | 2.65 (1.29, 5.92) | 0.05 (0.01, 0.31) | 0.09 (0.02, 0.48) | 0.0021 (0.0005, 0.0132) | 0.0101 (0.0019, 0.0543) | 5 |
| Brazil | 6.4 (2.2, 19.4) | 3.38 (1.65, 8.4) | 0.1 (0.02, 0.55) | 0.15 (0.03, 0.61) | 0.006 (0.0013, 0.0342) | 0.0174 (0.0035, 0.0772) | 5 |
| Brunei | 4.3 (1.5, 12.5) | 2.61 (0.79, 6.96) | 0.14 (0.02, 0.59) | 0.31 (0.03, 0.88) | 0.0084 (0.0014, 0.0378) | 0.0629 (0.0068, 0.2025) | 4 |
| Bulgaria | 5.9 (2.3, 15.5) | 5.15 (1.97, 13.29) | 0.08 (0.02, 0.38) | 0.18 (0.04, 0.61) | 0.0026 (0.0006, 0.0133) | 0.0203 (0.0043, 0.0745) | 4 |
| Burkina Faso | 4.9 (1.9, 13.1) | 4.03 (1.58, 11.53) | 0.09 (0.02, 0.42) | 0.17 (0.04, 0.61) | 0.0043 (0.001, 0.0223) | 0.0195 (0.0039, 0.0762) | 3 |
| Burma | 2.7 (1.3, 9.4) | 2.03 (0.88, 8.53) | 0.06 (0.01, 0.61) | 0.06 (0.0, 0.61) | 0.0111 (0.0019, 0.1166) | 0.0173 (0.0012, 0.2104) | 2 |
| Cabo Verde | 3.4 (1.2, 12.4) | 2.37 (0.7, 10.1) | 0.11 (0.02, 1.47) | 0.15 (0.01, 1.67) | 0.0221 (-0.0208, 0.3289) | 0.0525 (-0.0726, 0.733) | 2 |
| Cameroon | 5.7 (1.9, 18.2) | 4.33 (1.51, 14.69) | 0.11 (0.02, 0.6) | 0.21 (0.03, 0.73) | 0.0086 (0.0014, 0.0519) | 0.0349 (0.0046, 0.1586) | 3 |
| Chile | 4.2 (2.0, 9.1) | 3.94 (1.84, 8.94) | 0.05 (0.01, 0.27) | 0.11 (0.02, 0.49) | 0.0013 (0.0003, 0.0073) | 0.0065 (0.0013, 0.033) | 5 |
| China | 9.6 (4.1, 19.2) | 0.96 (0.38, 1.56) | 0.08 (0.03, 0.21) | 0.16 (0.02, 0.66) | 0.0007 (0.0002, 0.002) | 0.0094 (0.0012, 0.0387) | 11 |
| Colombia | 6.6 (2.5, 18.1) | 4.83 (1.81, 13.04) | 0.12 (0.03, 0.49) | 0.25 (0.06, 0.68) | 0.0031 (0.0007, 0.0137) | 0.0204 (0.0049, 0.0611) | 4 |
| Congo (Brazzaville) | 3.6 (1.4, 12.5) | 2.27 (0.7, 9.02) | 0.09 (0.01, 0.94) | 0.12 (0.01, 0.96) | 0.0154 (0.0018, 0.1975) | 0.036 (0.0016, 0.5313) | 3 |
| Congo (Kinshasa) | 4.1 (1.5, 11.3) | 2.86 (1.15, 9.25) | 0.09 (0.02, 0.61) | 0.15 (0.03, 0.69) | 0.0103 (0.0022, 0.0732) | 0.0407 (0.0064, 0.2076) | 3 |
| Costa Rica | 4.7 (1.8, 13.6) | 3.12 (1.31, 8.73) | 0.09 (0.02, 0.46) | 0.17 (0.03, 0.64) | 0.0037 (0.0008, 0.0215) | 0.0231 (0.0041, 0.0975) | 4 |
| Croatia | 3.1 (1.5, 7.1) | 2.65 (1.32, 6.89) | 0.04 (0.01, 0.28) | 0.07 (0.01, 0.54) | 0.0017 (0.0004, 0.0112) | 0.0058 (0.0006, 0.0498) | 6 |
| Cuba | 3.7 (1.6, 10.4) | 2.77 (1.28, 8.21) | 0.06 (0.02, 0.45) | 0.08 (0.02, 0.48) | 0.0027 (0.0006, 0.0207) | 0.0074 (0.0013, 0.047) | 4 |
| Cyprus | 6.5 (2.2, 18.0) | 3.73 (1.4, 10.95) | 0.14 (0.03, 0.57) | 0.27 (0.06, 0.74) | 0.0067 (0.0013, 0.0312) | 0.0405 (0.0076, 0.1334) | 4 |
| Czechia | 3.9 (1.8, 8.8) | 2.89 (1.39, 6.27) | 0.06 (0.02, 0.38) | 0.13 (0.03, 0.57) | 0.0016 (0.0004, 0.0108) | 0.0114 (0.0022, 0.0539) | 5 |
| Denmark | 6.0 (2.5, 15.4) | 3.6 (1.53, 8.47) | 0.06 (0.01, 0.34) | 0.18 (0.03, 0.63) | 0.0022 (0.0005, 0.0128) | 0.0194 (0.0035, 0.0765) | 5 |
| Diamond Princess | 3.1 (1.3, 8.3) | 0.29 (0.01, 0.85) | 0.09 (0.02, 0.68) | 0.2 (0.02, 0.88) | 0.0086 (0.0014, 0.0602) | 0.0614 (0.0052, 0.3591) | 9 |
| Djibouti | 4.2 (1.6, 13.7) | 2.54 (0.95, 8.5) | 0.08 (0.02, 0.53) | 0.09 (0.01, 0.57) | 0.0068 (0.0013, 0.049) | 0.012 (0.0014, 0.092) | 2 |
| Dominican Republic | 4.3 (1.9, 10.7) | 4.01 (1.7, 12.67) | 0.05 (0.01, 0.29) | 0.07 (0.01, 0.46) | 0.0031 (0.0006, 0.0204) | 0.0106 (0.0016, 0.0709) | 4 |
| Ecuador | 3.3 (1.5, 8.6) | 3.39 (1.48, 10.94) | 0.04 (0.01, 0.3) | 0.04 (0.0, 0.4) | 0.0032 (0.0006, 0.0253) | 0.0057 (0.0005, 0.0612) | 5 |
| Egypt | 4.9 (1.7, 13.7) | 2.11 (1.13, 6.06) | 0.11 (0.03, 0.62) | 0.18 (0.04, 0.68) | 0.0053 (0.0014, 0.0292) | 0.0213 (0.005, 0.0818) | 4 |
| El Salvador | 5.0 (1.8, 14.2) | 4.14 (1.49, 12.83) | 0.1 (0.02, 0.56) | 0.16 (0.03, 0.65) | 0.0069 (0.0014, 0.0406) | 0.0209 (0.0032, 0.0993) | 2 |
| Estonia | 4.6 (1.9, 12.8) | 3.55 (1.51, 9.52) | 0.07 (0.02, 0.38) | 0.15 (0.03, 0.57) | 0.0031 (0.0007, 0.0174) | 0.0198 (0.0037, 0.0859) | 4 |
| Ethiopia | 2.8 (1.3, 9.6) | 2.12 (0.93, 7.84) | 0.07 (0.01, 0.75) | 0.07 (0.01, 0.72) | 0.0109 (0.0019, 0.1224) | 0.0218 (0.0018, 0.2655) | 3 |
| Finland | 4.8 (1.9, 14.7) | 3.24 (1.53, 7.86) | 0.07 (0.02, 0.46) | 0.15 (0.03, 0.61) | 0.0047 (0.001, 0.0312) | 0.0249 (0.0041, 0.1203) | 5 |
| France | 1.5 (1.1, 5.1) | 1.21 (0.73, 2.55) | 0.53 (0.01, 7.18) | 0.11 (0.02, 0.63) | 0.0422 (0.001, 0.6573) | 0.0163 (0.003, 0.1028) | 10 |
| Gabon | 3.2 (1.3, 10.7) | 2.2 (0.8, 8.6) | 0.07 (0.01, 0.87) | 0.08 (0.01, 0.89) | 0.0129 (0.0019, 0.1629) | 0.023 (0.0016, 0.354) | 2 |
| Georgia | 4.6 (1.7, 12.5) | 1.81 (1.11, 4.45) | 0.09 (0.02, 0.57) | 0.16 (0.03, 0.65) | 0.0056 (0.0013, 0.0364) | 0.0238 (0.0039, 0.1085) | 5 |
| Germany | 1.7 (1.0, 4.9) | 0.9 (0.56, 1.79) | 0.19 (0.01, 8.09) | 0.11 (0.03, 0.71) | 0.0074 (0.0004, 0.3351) | 0.0081 (0.0018, 0.0614) | 10 |
| Ghana | 5.2 (1.7, 15.8) | 3.69 (1.41, 12.56) | 0.1 (0.02, 0.61) | 0.17 (0.03, 0.71) | 0.0105 (0.0019, 0.072) | 0.0361 (0.0039, 0.2002) | 3 |
| Greece | 4.4 (2.0, 12.0) | 2.76 (1.4, 5.82) | 0.06 (0.02, 0.4) | 0.19 (0.03, 0.67) | 0.0038 (0.0008, 0.0247) | 0.0407 (0.0067, 0.1635) | 6 |
| Guatemala | 3.2 (1.3, 10.7) | 2.29 (1.03, 8.5) | 0.07 (0.02, 0.61) | 0.09 (0.01, 0.61) | 0.0091 (0.0018, 0.0768) | 0.0208 (0.002, 0.1621) | 3 |
| Guinea | 4.4 (1.7, 14.3) | 3.51 (1.31, 12.58) | 0.08 (0.02, 0.55) | 0.1 (0.01, 0.6) | 0.0076 (0.0014, 0.0553) | 0.0153 (0.0018, 0.1137) | 2 |
| Guyana | 3.5 (1.2, 12.0) | 2.22 (0.64, 9.12) | 0.16 (0.02, 1.34) | 0.25 (-0.07, 1.49) | 0.0318 (-0.0086, 0.3779) | 0.1119 (-0.0393, 0.8531) | 3 |
| Haiti | 3.6 (1.2, 12.6) | 2.88 (0.81, 11.47) | 0.15 (0.02, 1.18) | 0.24 (0.01, 1.25) | 0.0308 (0.0019, 0.3536) | 0.0929 (-0.0005, 0.7396) | 2 |
| Honduras | 3.9 (1.6, 12.7) | 2.85 (1.24, 8.47) | 0.07 (0.02, 0.48) | 0.13 (0.02, 0.62) | 0.0089 (0.0016, 0.0672) | 0.0384 (0.0052, 0.2132) | 4 |
| Hungary | 5.0 (2.0, 14.0) | 3.23 (1.49, 7.81) | 0.07 (0.02, 0.42) | 0.14 (0.03, 0.55) | 0.0035 (0.0008, 0.0203) | 0.0177 (0.0035, 0.0742) | 5 |
| Iceland | 4.4 (1.8, 14.5) | 2.58 (1.34, 7.08) | 0.11 (0.02, 0.48) | 0.31 (0.06, 0.77) | 0.0026 (0.0006, 0.012) | 0.0098 (0.0017, 0.0419) | 5 |
| India | 6.8 (2.3, 23.6) | 2.64 (1.5, 5.17) | 0.14 (0.04, 0.67) | 0.22 (0.05, 0.7) | 0.004 (0.001, 0.0194) | 0.014 (0.0034, 0.0484) | 5 |
| Indonesia | 5.6 (2.3, 14.5) | 5.21 (2.1, 14.94) | 0.05 (0.01, 0.31) | 0.12 (0.02, 0.6) | 0.0021 (0.0005, 0.0133) | 0.0159 (0.0031, 0.0845) | 5 |
| Iran | 7.9 (3.2, 17.4) | 1.34 (1.1, 1.86) | 0.06 (0.02, 0.28) | 0.29 (0.07, 0.73) | 0.0016 (0.0004, 0.0075) | 0.0272 (0.0062, 0.0722) | 7 |
| Iraq | 2.5 (1.3, 9.6) | 1.65 (1.11, 3.18) | 0.09 (0.02, 0.79) | 0.2 (0.03, 0.8) | 0.0036 (0.0008, 0.0295) | 0.0188 (0.0027, 0.0789) | 6 |
| Ireland | 5.5 (2.0, 16.6) | 3.2 (1.53, 7.74) | 0.08 (0.02, 0.51) | 0.14 (0.03, 0.6) | 0.0055 (0.0012, 0.0354) | 0.0229 (0.0042, 0.1075) | 5 |
| Israel | 3.7 (1.8, 8.3) | 3.05 (1.54, 6.34) | 0.05 (0.02, 0.34) | 0.1 (0.02, 0.48) | 0.0013 (0.0003, 0.0083) | 0.0053 (0.0011, 0.0284) | 5 |
| Italy | 3.4 (1.0, 8.4) | 0.83 (0.21, 1.19) | 0.05 (0.01, 5.16) | 0.03 (0.01, 0.54) | 0.0031 (0.0005, 0.3228) | 0.0062 (0.0014, 0.1103) | 10 |
| Jamaica | 2.2 (1.2, 6.2) | 0.94 (0.5, 3.55) | 0.05 (0.01, 0.46) | 0.04 (0.01, 0.4) | 0.0091 (0.0018, 0.0839) | 0.0121 (0.0012, 0.1249) | 3 |
| Japan | 3.3 (1.5, 7.2) | 1.93 (1.25, 3.36) | 0.05 (0.02, 0.41) | 0.08 (0.01, 0.44) | 0.0023 (0.0006, 0.0168) | 0.0081 (0.0013, 0.0481) | 10 |
| Jordan | 6.1 (2.2, 16.9) | 5.31 (1.91, 15.22) | 0.12 (0.03, 0.48) | 0.28 (0.06, 0.73) | 0.0049 (0.0011, 0.022) | 0.0274 (0.0054, 0.0855) | 3 |
| Kazakhstan | 5.4 (2.0, 14.4) | 3.14 (1.36, 9.36) | 0.1 (0.03, 0.53) | 0.16 (0.04, 0.61) | 0.0034 (0.0009, 0.0177) | 0.0131 (0.0029, 0.0526) | 4 |
| Kenya | 4.2 (1.7, 12.9) | 3.45 (1.41, 10.74) | 0.08 (0.02, 0.5) | 0.13 (0.02, 0.61) | 0.0057 (0.0011, 0.0397) | 0.0203 (0.0028, 0.1114) | 3 |
| Korea, South | 3.1 (1.4, 5.8) | 0.43 (0.05, 0.87) | 0.04 (0.01, 0.79) | 0.1 (0.0, 0.74) | 0.0014 (0.0003, 0.0287) | 0.0071 (0.0002, 0.066) | 9 |
| Kosovo | 2.4 (1.2, 8.0) | 1.88 (0.91, 7.16) | 0.06 (0.02, 0.47) | 0.06 (0.01, 0.5) | 0.0076 (0.0015, 0.0671) | 0.0143 (0.0017, 0.1404) | 2 |
| Kuwait | 4.3 (1.4, 15.9) | 1.79 (1.15, 3.5) | 0.07 (0.02, 1.09) | 0.09 (0.01, 0.73) | 0.0037 (0.0009, 0.0506) | 0.01 (0.0007, 0.0893) | 6 |
| Kyrgyzstan | 5.2 (1.8, 15.2) | 3.71 (1.44, 12.44) | 0.11 (0.02, 0.6) | 0.17 (0.03, 0.68) | 0.0075 (0.0015, 0.0452) | 0.026 (0.0041, 0.1207) | 3 |
| Latvia | 4.7 (1.8, 14.4) | 3.06 (1.26, 9.05) | 0.09 (0.02, 0.53) | 0.19 (0.03, 0.71) | 0.007 (0.0013, 0.0465) | 0.0406 (0.0058, 0.1909) | 4 |
| Lebanon | 4.3 (1.9, 13.3) | 2.53 (1.2, 5.94) | 0.07 (0.02, 0.49) | 0.2 (0.03, 0.71) | 0.0033 (0.0007, 0.0221) | 0.0321 (0.005, 0.1285) | 5 |
| Liberia | 4.9 (1.6, 15.3) | 4.3 (1.27, 14.09) | 0.12 (0.02, 0.8) | 0.18 (0.02, 0.87) | 0.0189 (0.0025, 0.1474) | 0.0427 (0.0032, 0.3093) | 1 |
| Libya | 3.9 (1.3, 13.4) | 3.18 (0.99, 12.24) | 0.1 (0.02, 1.01) | 0.13 (0.01, 1.11) | 0.0178 (0.0012, 0.2248) | 0.0331 (0.0017, 0.4473) | 1 |
| Liechtenstein | 4.6 (1.3, 15.4) | 2.66 (0.62, 10.71) | 0.21 (0.02, 1.31) | 0.38 (-0.05, 1.51) | 0.0332 (-0.0119, 0.3348) | 0.1426 (-0.0647, 0.841) | 3 |
| Lithuania | 5.3 (2.1, 15.5) | 4.22 (1.67, 11.85) | 0.08 (0.02, 0.44) | 0.19 (0.04, 0.66) | 0.0038 (0.0008, 0.0241) | 0.0288 (0.0051, 0.1178) | 4 |
| Luxembourg | 6.1 (2.5, 15.9) | 4.69 (1.85, 12.82) | 0.08 (0.02, 0.43) | 0.3 (0.06, 0.78) | 0.0038 (0.0008, 0.0223) | 0.0535 (0.0096, 0.1865) | 4 |
| Malaysia | 1.8 (1.2, 3.4) | 1.12 (0.72, 1.76) | 0.03 (0.01, 0.17) | 0.2 (0.05, 0.46) | 0.0015 (0.0005, 0.0074) | 0.0111 (0.0027, 0.0305) | 10 |
| Mali | 7.1 (2.2, 19.7) | 5.93 (1.99, 18.52) | 0.15 (0.04, 0.67) | 0.25 (0.06, 0.76) | 0.0124 (0.0027, 0.0606) | 0.0409 (0.0077, 0.1521) | 2 |
| Malta | 4.5 (1.7, 12.9) | 2.43 (1.15, 7.18) | 0.09 (0.02, 0.6) | 0.16 (0.03, 0.69) | 0.0081 (0.0016, 0.056) | 0.0344 (0.0049, 0.1712) | 4 |
| Mauritius | 5.8 (1.8, 17.2) | 4.4 (1.46, 14.06) | 0.13 (0.02, 0.59) | 0.26 (0.04, 0.77) | 0.0086 (0.0015, 0.0455) | 0.0464 (0.007, 0.1733) | 3 |
| Mexico | 3.0 (1.4, 8.4) | 2.95 (1.4, 6.71) | 0.05 (0.01, 0.35) | 0.04 (0.01, 0.3) | 0.0038 (0.0009, 0.0307) | 0.0045 (0.0007, 0.0365) | 5 |
| Moldova | 6.1 (2.2, 17.6) | 3.79 (1.58, 10.86) | 0.11 (0.03, 0.53) | 0.17 (0.04, 0.62) | 0.0034 (0.0008, 0.0179) | 0.0138 (0.003, 0.0535) | 4 |
| Monaco | 4.4 (1.4, 12.7) | 3.21 (1.02, 10.28) | 0.1 (0.02, 0.82) | 0.19 (0.02, 0.91) | 0.0155 (0.0027, 0.1382) | 0.0668 (0.0067, 0.3948) | 3 |
| Montenegro | 6.3 (2.1, 17.9) | 4.55 (1.59, 14.45) | 0.14 (0.03, 0.63) | 0.26 (0.05, 0.77) | 0.0104 (0.0018, 0.0538) | 0.0487 (0.0079, 0.1812) | 3 |
| Morocco | 6.3 (2.5, 15.5) | 5.69 (2.25, 14.53) | 0.08 (0.02, 0.35) | 0.21 (0.05, 0.64) | 0.0024 (0.0006, 0.011) | 0.022 (0.0052, 0.0715) | 4 |
| Netherlands | 5.2 (2.3, 13.2) | 4.15 (1.9, 9.34) | 0.06 (0.02, 0.31) | 0.17 (0.04, 0.6) | 0.0062 (0.0013, 0.0558) | 0.0498 (0.0094, 0.2306) | 5 |
| New Zealand | 3.3 (1.5, 8.3) | 3.63 (1.55, 12.14) | 0.03 (0.01, 0.28) | 0.03 (0.0, 0.58) | 0.0015 (0.0003, 0.0124) | 0.002 (0.0001, 0.0403) | 5 |
| Niger | 5.0 (1.7, 15.6) | 3.81 (1.26, 12.6) | 0.09 (0.02, 0.51) | 0.14 (0.02, 0.66) | 0.0069 (0.0011, 0.0454) | 0.0216 (0.0025, 0.1255) | 2 |
| Nigeria | 5.4 (2.0, 13.9) | 4.04 (1.55, 11.73) | 0.11 (0.03, 0.48) | 0.18 (0.04, 0.62) | 0.0046 (0.0011, 0.0221) | 0.0183 (0.0039, 0.0684) | 3 |
| North Macedonia | 4.6 (1.9, 13.3) | 3.47 (1.55, 9.41) | 0.07 (0.02, 0.46) | 0.12 (0.02, 0.56) | 0.0039 (0.0008, 0.0255) | 0.0163 (0.0027, 0.0827) | 4 |
| Norway | 5.2 (2.2, 14.1) | 3.4 (1.53, 7.46) | 0.07 (0.02, 0.36) | 0.21 (0.04, 0.68) | 0.0051 (0.0009, 0.1817) | 0.0542 (0.009, 0.4432) | 5 |
| Oman | 2.6 (1.3, 7.9) | 2.28 (1.29, 4.58) | 0.05 (0.01, 0.38) | 0.04 (0.01, 0.3) | 0.0043 (0.001, 0.0342) | 0.0054 (0.0008, 0.0463) | 6 |
| Pakistan | 3.5 (1.6, 8.5) | 3.36 (1.59, 9.15) | 0.04 (0.01, 0.28) | 0.07 (0.01, 0.47) | 0.0019 (0.0004, 0.0121) | 0.0066 (0.0006, 0.0461) | 6 |
| Panama | 5.4 (2.0, 15.9) | 3.38 (1.41, 10.05) | 0.1 (0.02, 0.49) | 0.17 (0.04, 0.61) | 0.0049 (0.0011, 0.026) | 0.0232 (0.0049, 0.0938) | 4 |
| Paraguay | 3.5 (1.4, 10.6) | 2.3 (1.12, 7.34) | 0.07 (0.02, 0.55) | 0.11 (0.02, 0.61) | 0.0076 (0.0015, 0.0594) | 0.0268 (0.0034, 0.1633) | 4 |
| Peru | 4.5 (1.8, 12.9) | 3.63 (1.66, 9.75) | 0.07 (0.02, 0.41) | 0.09 (0.02, 0.46) | 0.0029 (0.0007, 0.0187) | 0.0065 (0.0013, 0.0355) | 4 |
| Philippines | 5.7 (2.1, 15.8) | 2.8 (1.09, 8.22) | 0.17 (0.05, 0.57) | 0.34 (0.09, 0.75) | 0.0059 (0.0015, 0.0207) | 0.0373 (0.0095, 0.0933) | 4 |
| Poland | 4.8 (1.9, 13.5) | 2.53 (1.05, 7.23) | 0.1 (0.03, 0.51) | 0.19 (0.05, 0.64) | 0.0031 (0.0008, 0.0158) | 0.0156 (0.0035, 0.0563) | 4 |
| Portugal | 4.9 (2.2, 10.5) | 4.33 (1.99, 9.17) | 0.05 (0.02, 0.31) | 0.14 (0.03, 0.57) | 0.0022 (0.0006, 0.0129) | 0.0172 (0.0037, 0.0761) | 5 |
| Qatar | 7.1 (2.4, 19.4) | 2.29 (1.22, 6.02) | 0.14 (0.03, 0.57) | 0.26 (0.06, 0.72) | 0.0055 (0.0012, 0.0257) | 0.0302 (0.0056, 0.1028) | 5 |
| Romania | 4.1 (2.0, 8.3) | 4.09 (1.98, 8.43) | 0.05 (0.01, 0.25) | 0.08 (0.02, 0.43) | 0.0014 (0.0004, 0.0075) | 0.0057 (0.0013, 0.0301) | 5 |
| Russia | 1.9 (1.1, 4.6) | 2.74 (1.33, 6.82) | 0.04 (0.01, 0.76) | 0.0 (0.0, 0.13) | 0.0021 (0.0005, 0.0391) | 0.0003 (0.0, 0.0084) | 9 |
| San Marino | 4.5 (1.6, 12.1) | 1.38 (0.85, 3.72) | 0.1 (0.02, 0.61) | 0.2 (0.04, 0.74) | 0.0112 (0.0023, 0.0711) | 0.0633 (0.0105, 0.2618) | 5 |
| Saudi Arabia | 4.9 (2.0, 12.3) | 4.17 (1.83, 9.86) | 0.06 (0.02, 0.32) | 0.12 (0.03, 0.5) | 0.0015 (0.0004, 0.0084) | 0.0079 (0.0016, 0.0354) | 5 |
| Senegal | 3.4 (1.7, 7.6) | 3.25 (1.64, 7.42) | 0.05 (0.01, 0.28) | 0.09 (0.02, 0.46) | 0.0028 (0.0007, 0.0167) | 0.009 (0.0016, 0.0477) | 5 |
| Serbia | 5.9 (2.1, 17.3) | 3.13 (1.36, 9.61) | 0.12 (0.03, 0.61) | 0.19 (0.04, 0.68) | 0.0077 (0.0017, 0.0438) | 0.0295 (0.0058, 0.1247) | 4 |
| Singapore | 3.8 (1.6, 8.7) | 2.27 (1.17, 4.72) | 0.06 (0.02, 0.43) | 0.01 (0.0, 0.32) | 0.002 (0.0005, 0.0152) | 0.0005 (0.0, 0.0157) | 11 |
| Slovakia | 7.0 (2.2, 19.2) | 3.25 (1.33, 10.2) | 0.15 (0.04, 0.65) | 0.28 (0.06, 0.78) | 0.0092 (0.0019, 0.0456) | 0.0453 (0.0086, 0.1595) | 4 |
| Slovenia | 7.0 (2.6, 19.4) | 4.02 (1.59, 9.6) | 0.12 (0.03, 0.5) | 0.33 (0.08, 0.78) | 0.0047 (0.0009, 0.0225) | 0.0592 (0.0128, 0.1663) | 5 |
| Somalia | 3.7 (1.5, 12.2) | 2.78 (1.06, 10.19) | 0.06 (0.01, 0.51) | 0.06 (0.01, 0.55) | 0.0105 (0.0016, 0.0958) | 0.0133 (0.0012, 0.1486) | 1 |
| South Africa | 5.8 (2.3, 17.5) | 5.01 (1.98, 13.49) | 0.08 (0.02, 0.45) | 0.19 (0.04, 0.66) | 0.0044 (0.0009, 0.0273) | 0.0285 (0.005, 0.1202) | 4 |
| Spain | 5.2 (2.5, 9.8) | 4.14 (2.1, 7.65) | 0.05 (0.02, 0.24) | 0.21 (0.05, 0.64) | 0.0017 (0.0005, 0.0079) | 0.0289 (0.0067, 0.0913) | 6 |
| Sri Lanka | 7.0 (2.4, 17.5) | 3.11 (1.04, 10.37) | 0.2 (0.05, 0.62) | 0.4 (0.1, 0.82) | 0.0103 (0.0026, 0.0355) | 0.0689 (0.0166, 0.1615) | 4 |
| Sudan | 3.4 (1.2, 11.8) | 3.2 (1.03, 11.68) | 0.11 (0.02, 1.05) | 0.13 (0.01, 1.13) | 0.022 (0.0018, 0.2961) | 0.0414 (0.0013, 0.5452) | 1 |
| Sweden | 5.2 (2.1, 14.4) | 2.27 (1.25, 5.1) | 0.08 (0.02, 0.42) | 0.17 (0.04, 0.6) | 0.0052 (0.0011, 0.0298) | 0.0334 (0.0065, 0.1377) | 6 |
| Switzerland | 3.6 (1.8, 8.0) | 3.12 (1.68, 6.5) | 0.06 (0.02, 0.33) | 0.16 (0.03, 0.68) | 0.002 (0.0005, 0.0124) | 0.0186 (0.0034, 0.0901) | 6 |
| Syria | 3.2 (1.2, 10.9) | 2.57 (0.77, 10.07) | 0.15 (0.02, 1.42) | 0.22 (-0.01, 1.49) | 0.0312 (-0.0053, 0.3978) | 0.0896 (-0.0146, 0.8161) | 2 |
| Taiwan* | 2.0 (1.2, 5.1) | 0.74 (0.18, 2.0) | 0.05 (0.01, 0.32) | 0.08 (0.0, 0.48) | 0.0045 (0.0011, 0.0324) | 0.0148 (0.0006, 0.0936) | 10 |
| Tanzania | 2.7 (1.3, 8.8) | 1.48 (0.65, 5.67) | 0.06 (0.01, 0.57) | 0.05 (0.0, 0.58) | 0.011 (0.0019, 0.1148) | 0.0145 (0.0009, 0.2131) | 3 |
| Thailand | 1.9 (1.2, 3.5) | 0.81 (0.18, 1.59) | 0.04 (0.01, 0.16) | 0.23 (0.04, 0.56) | 0.0024 (0.0007, 0.0107) | 0.016 (0.0027, 0.0556) | 11 |
| Togo | 4.1 (1.4, 11.4) | 2.94 (1.03, 9.48) | 0.09 (0.02, 0.73) | 0.17 (0.02, 0.82) | 0.0113 (0.0023, 0.09) | 0.0431 (0.0048, 0.2472) | 3 |
| Trinidad and Tobago | 5.6 (1.9, 17.9) | 4.3 (1.37, 12.93) | 0.14 (0.02, 0.61) | 0.32 (0.04, 0.86) | 0.0156 (0.0022, 0.0917) | 0.1051 (0.0128, 0.3855) | 3 |
| Tunisia | 4.7 (1.9, 14.2) | 3.43 (1.45, 9.6) | 0.08 (0.02, 0.47) | 0.16 (0.03, 0.64) | 0.0056 (0.0011, 0.0359) | 0.033 (0.0053, 0.1501) | 4 |
| Turkey | 8.2 (3.2, 20.1) | 8.45 (3.27, 25.28) | 0.05 (0.01, 0.26) | 0.13 (0.03, 0.51) | 0.0009 (0.0002, 0.0046) | 0.0068 (0.0014, 0.0267) | 3 |
| Ukraine | 6.1 (2.4, 16.8) | 5.79 (2.28, 16.3) | 0.07 (0.02, 0.37) | 0.12 (0.03, 0.5) | 0.0024 (0.0005, 0.0135) | 0.0097 (0.002, 0.0437) | 3 |
| United Arab Emirates | 1.8 (1.1, 4.1) | 2.22 (1.23, 5.1) | 0.04 (0.01, 0.48) | 0.01 (0.0, 0.18) | 0.0035 (0.0008, 0.0409) | 0.0011 (0.0001, 0.0204) | 10 |
| United Kingdom | 2.7 (1.3, 6.3) | 2.61 (1.31, 6.58) | 0.04 (0.01, 0.33) | 0.01 (0.0, 0.3) | 0.0061 (0.0013, 0.0649) | 0.0021 (0.0001, 0.0822) | 9 |
| Uruguay | 6.2 (2.1, 18.8) | 3.44 (1.13, 11.59) | 0.13 (0.03, 0.54) | 0.29 (0.05, 0.77) | 0.0072 (0.0013, 0.0358) | 0.0388 (0.0063, 0.139) | 3 |
| Uzbekistan | 4.4 (1.6, 13.7) | 3.07 (1.31, 10.6) | 0.08 (0.02, 0.62) | 0.11 (0.02, 0.62) | 0.005 (0.0012, 0.0368) | 0.0134 (0.0021, 0.08) | 3 |
| Venezuela | 8.4 (2.7, 23.2) | 5.69 (1.86, 17.88) | 0.23 (0.05, 0.68) | 0.47 (0.11, 0.87) | 0.0153 (0.0031, 0.0555) | 0.0804 (0.0171, 0.2025) | 3 |
| **West Bank and Gaza** | **3.9 (1.4, 12.0)** | **1.6 (0.99, 3.97)** | **0.09 (0.02, 0.68)** | **0.14 (0.01, 0.7)** | **0.0087 (0.0018, 0.0688)** | **0.0312 (0.0027, 0.1843)** | **5** |
| **Zambia** | **4.0 (1.2, 14.4)** | **2.79 (0.7, 11.78)** | **0.16 (0.02, 1.19)** | **0.29 (-0.04, 1.31)** | **0.0295 (-0.001, 0.3355)** | **0.1194 (-0.0155, 0.7793)** | **3** |
| **Zimbabwe** | **3.5 (1.2, 12.0)** | **2.69 (0.79, 10.72)** | **0.17 (0.02, 1.29)** | **0.25 (-0.01, 1.39)** | **0.0345 (-0.0033, 0.3862)** | **0.1018 (-0.0159, 0.7856)** | **2** |

**Figure S2.**
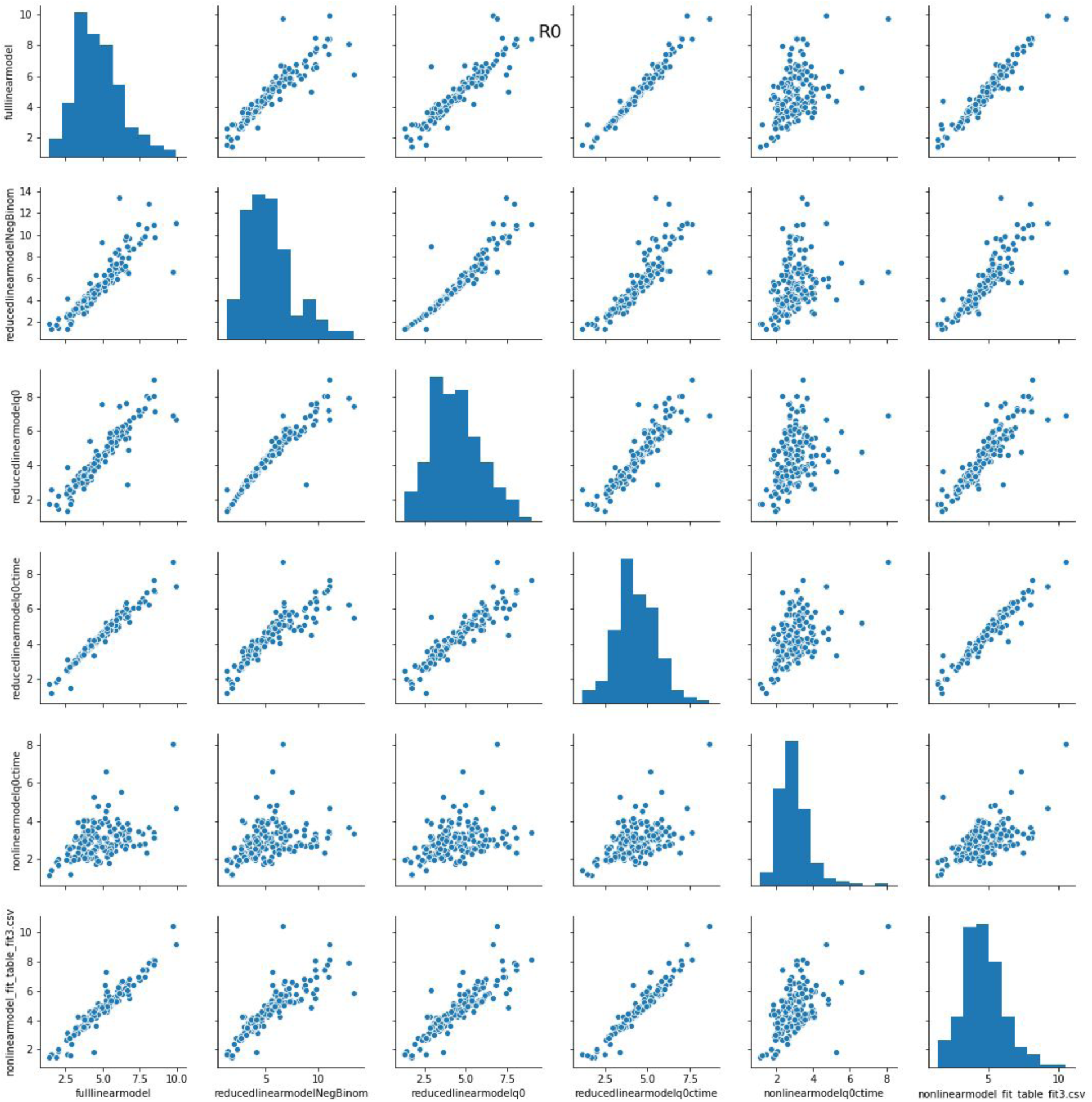
R_0_ estimates across models showing model consistency.

**Figure S3.**
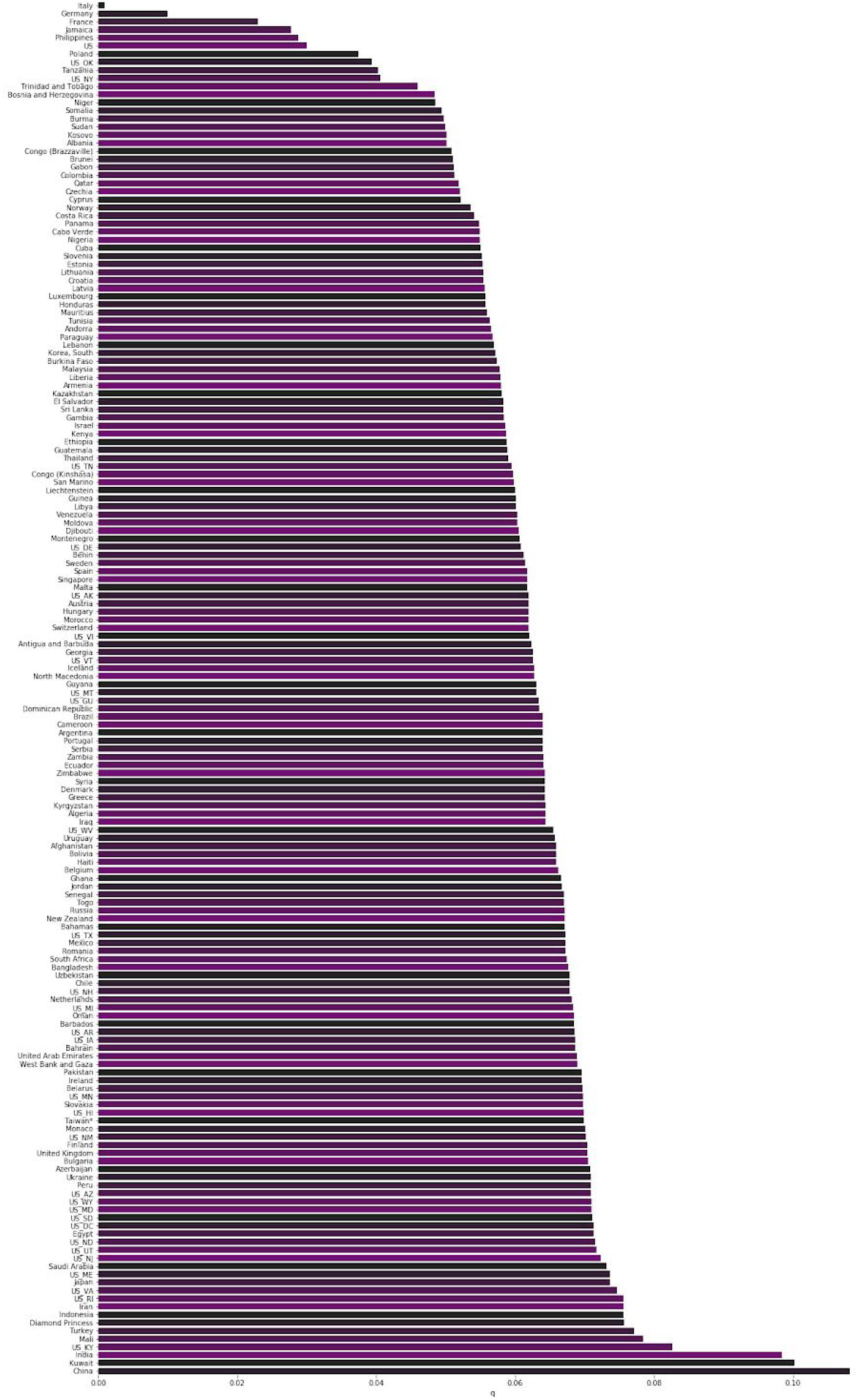
Median case infectiousness factor q for all regions.

**Figure S4.**
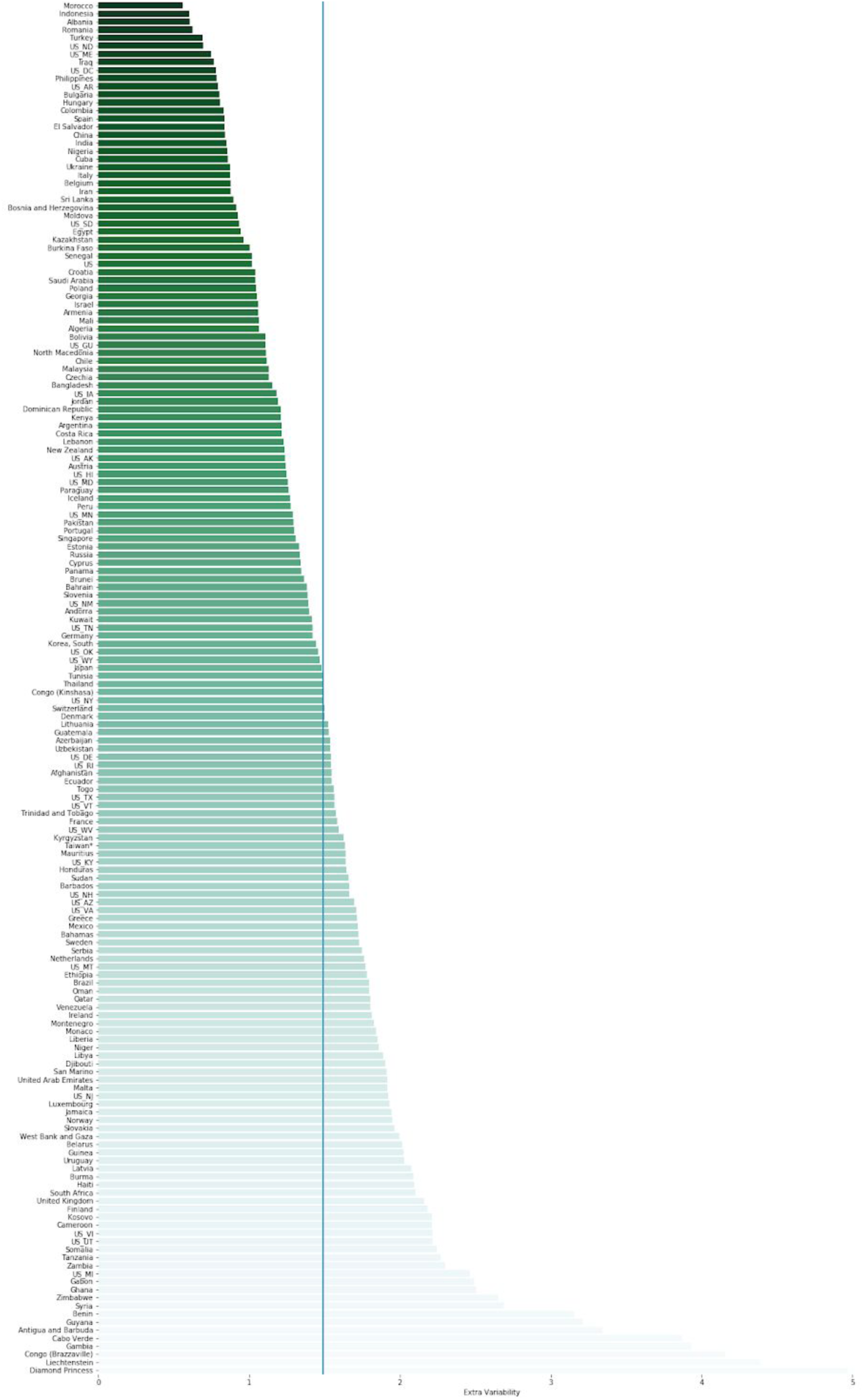
Median overdispersion factor for all regions. Blue line is median of 1.5 across all regions.

## S1 Latent variable SIR Model

We consider a latent variable SIR model where the observables are cases *C*, case recoveries *R_C_*, and case deaths *D_C_*. The latent variables are the numbers of susceptible *S* and undetected infected *I* individuals. Assuming probability of contact between people is completely random and uniform and ignoring the possibility of births, migration, or deaths due to other causes, the mass action equations for the model are

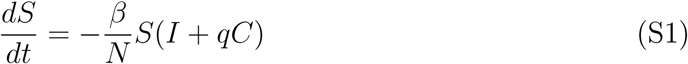

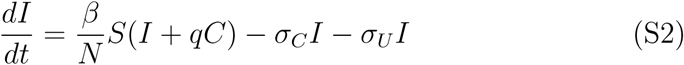

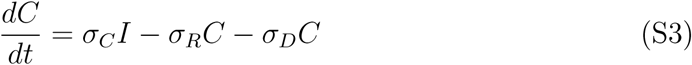

where *N* is the population of the region in question, *β* is the infection rate per contact, *σ_C_* is the transition rate from *I* to *C*, *σ_U_* is the disappearance rate of *I* not transitioning to *C*, and *σ* = *σ_R_* + *σ_D_* is the disappearance rate for cases. The parameter *q* accounts for possible differences in virus transmission rate from non-cases. It can be less than one due to say lower social contact or higher due to a larger viral load. If *q* = 0 then the model is identical to an SEIR model.

The equations obey the conservation condition

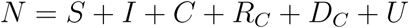

where

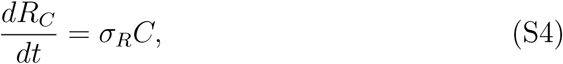

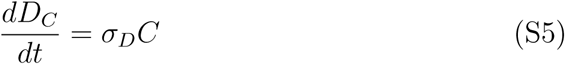

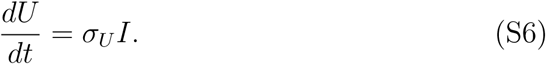

The population *U* represents the portion of *I* that either recover or die but remain undetected.

If initially *S* = *N*, *I* << *N*, and the parameters are such that the right hand side of (S2) is positive, then *I* will grow until it reaches a peak where 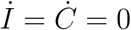, which is given by the condition

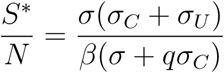

where *σ* = *σ_R_* + *σ_D_*. The fraction of the population remaining susceptible at the peak of the epidemic is the inverse of the reproduction number

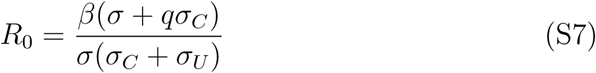

(*R*_0_ can also be computed using the next generation method). The pandemic will spread if *R*_0_ > 1. The total number that becomes infected and thus are no longer susceptible is *N_I_* = *N* − *S*. The peak of this number is given by

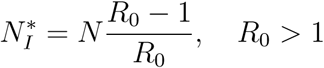

*I* will decrease after *N_I_* passes 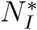. Thus, the pandemic can be mitigated by reducing 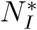, which can be achieved by either decreasing *R*_0_ or decreasing *N*. We model the effects of mitigation with a time dependence in *β* via *β → β_t_* where

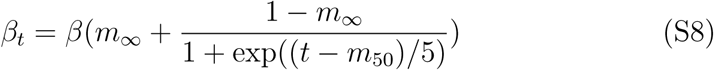

where *t* = 0 is the day of the first case. *β_t_* transitions from its initial value to a new value *βm*_∞_ on day *m*_50_ with a transition time of 5 days. We also account for the possibility that the case detection rate can change in time with with *σ_C_ → σt* where

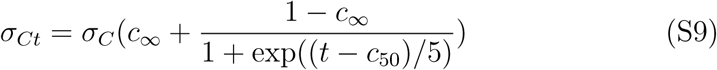

For example, the method of classification of cases changed in China on a single day leading to spike in the number of cases and this time dependence accounts for situations such as that.

The mean field SIR model assumes a well mixed population where the probability of interactions are homogeneous that does not reflect actual human interactions. An actual epidemic, rather than growing homogeneously throughout the population, will be seeded locally and spread within clusters that then propagate to other local clusters. Each cluster may locally saturate before spreading to another cluster and thus the average dynamics of the epidemic over a large region is not reflected by the actual spread at the local level. Hence, the population size *N* in the SIR model is not simply the regional population but a complex aggregation over many interacting local clusters, which may be difficult to estimate. This is a major limitation of mean field models since *N* is a major factor in determining the eventual rise and fall of the pandemic. However, this limitation can be circumvented by noting that in the initial stages of the epidemic where *I* is small and less than even the cluster population, we can fix *S* = *N* and the system (S1), (S2), and (S3) reduces to the two dimensional linear system

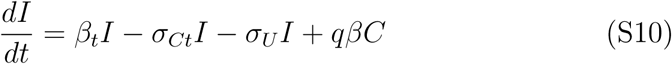

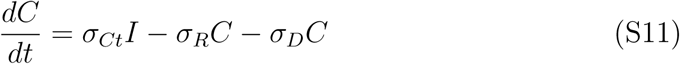

*N* is scaled out of the dynamics.

The eigenvalues for the linear system (S10) and (S11) assuming that *β_t_* and *σ_Ct_* are constant in time are

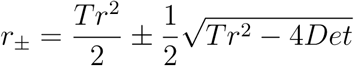

where *Tr* = *β_t_* − *σ_Ct_* − *σ_U_* − *σ* and Det = −*σ*(*β_t_* − *σ_Ct_* − *σ_U_*) − *qβσ_Ct_*. For *q* = 0 the eigenvalues simplify to

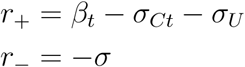

For most parameter values *r*_+_ will dominate the temporal dynamics. The pandemic will grow when it is positive and begin to extinguish only when *β_t_* falls below *σ_Ct_* + *σ_U_*.

We can use the eigenvalues for a lower bound on the rate for the pandemic to extinguish. In the nonlinear equations, we can consider an effective infectiousness *β*\* = *β_t_S*(*t*)/*N* = *β_t_*(1 − *n_I_*), where *n_I_* is the fraction of the population that is immune, either innately or by recovering from the infection.

From (S7) we can set

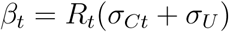

for *q* = 0 to obtain

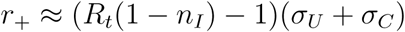

Thus the pandemic will extinguish at the rate of infection disappearance moderated by the residual spread.

## S2 Parameter identifiability

Here, we investigate whether the parameters in (*S*10) and (*S*11) are in principle identifiable from the observable data. We first consider the case without mitigation in which there are six parameters, *β*, *σ_C_*, *σ_R_*, *σ_D_*, *σ_U_*, and *q* together with the dynamic variables *I*(*t*) and *C*(*t*). The observable data consists of the time series of cases, case recoveries, and case deaths, which we denote by *λ_C_*(*t*), *λ_R_*(*t*),and *λ_D_*(*t*), respectively. For this analysis, we assume perfect information with no uncertainty.

From the data, we can immediately construct the following conditions

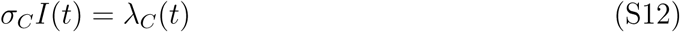

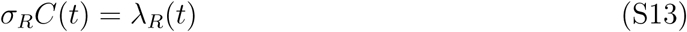

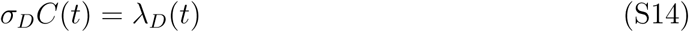

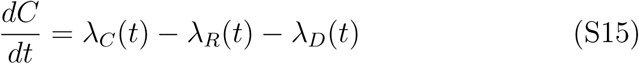

From these conditions we immediately obtain *C*(*t*), *σ_R_*, and *σ_D_*. Dividing (S10) by (S11), we obtain

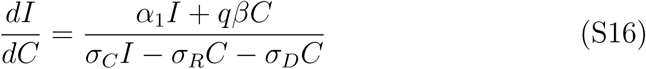

where *α*_1_ = *β* − *σ_U_* − *σ_C_*. Given that the epidemic is initiated with *I* > 0 and *C* = 0, then prior to the first case arising we can assume *dI/dC* = *α*_1_/*σ_C_*, from.which we can integrate to obtain *σ_C_I*(0) = *α*_1_*C*(0), where *t* = 0 marks the day of appearance of the first case. This then gives *α*_1_ = *λ_C_*(0)/*C*(0).

Multiplying (*S*10) by *σ_C_* gives

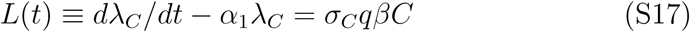

and thus *σ_C_qβ* = *L/C*. Finally, we can also rewrite (S10) as

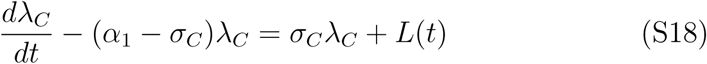

from which *σ_C_* can be inferred. Given *σ_C_* we can then infer *I*(*t*) from *λ_C_*(*t*) from which we can derive conditions for *β* − *σ_U_* and *qβ*, which are only two conditions for three unknowns. Thus, the observable data can at most specify five out of the six unknown parameters.

We can resolve this nonidentifiability in two ways. The first is by specifying one of the parameters. For example, we can specify *σ_U_* given that we have a prior on the rate of recovery or death from the infection. We can also specify *q* since it is a measure of how effective the isolation of cases are. The second is to utilize the fact that mitigation acts as a time dependent perturbation on *β* assuming all the other parameters are fixed. The mitigation is set by two parameters, the day it is applied and the effectiveness in diminishing *β*. Essentially the model can be applied separately before and after mitigation where only *β* has a different value. If the day of mitigation is known then this results in four conditions for four unknowns, i.e. *β_before_*, *β_after_*, *q*, and *σ_U_* and the parameters can be identified. Thus in principle, with perfect information, the model can be identified if given some prior information.

## S3 Parameter estimation

We estimate the parameters and their uncertainties using Bayesian methods, by putting prior distributions on each of the model parameters. A schematic of our overall Bayesian latent-variable model is shown in Fig. S1

We consider various versions of the model with varying number of parameters and weigh them using model comparison measures such as WAIC and LOO.

Considering the mean field SIR equations as a birth and death process implies a Poisson likelihood. However, in addition to this inherent stochasticity, errors will be introduced due to variability in criteria for measurement and recording. We account for this additional variance with a Negative Binomial distribution cf.

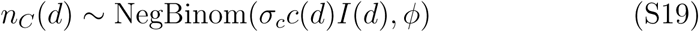

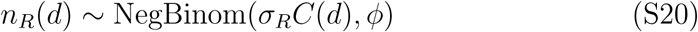

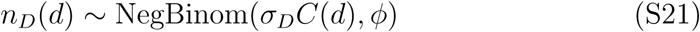

where *n_C,R,D_* are the total number of new cases, recovered cases, and case deaths on day *d*, *I*(*d*), *C*(*d*) are the total number for day *d*, and *ϕ* is a fitted factor quantifying the extra variance where *x* ~ NegBinom(*μ*, *ϕ*) with *E*[*X*] = *μ* and var[X] = *μ* + *μ*^2^/*ϕ*. We use inverse gamma distributed priors on *σ_U_*, *σ_R_* and *σ_D_* using parameters derived from ref X. We fit from the first day in which the daily case count is greater than one, which we call day 0. We then set *C*(0) to this count and *I*(0) = (*β* − *σ_U_* − *σ_C_*)/*σ_C_C*(0).

**Figure S1:**
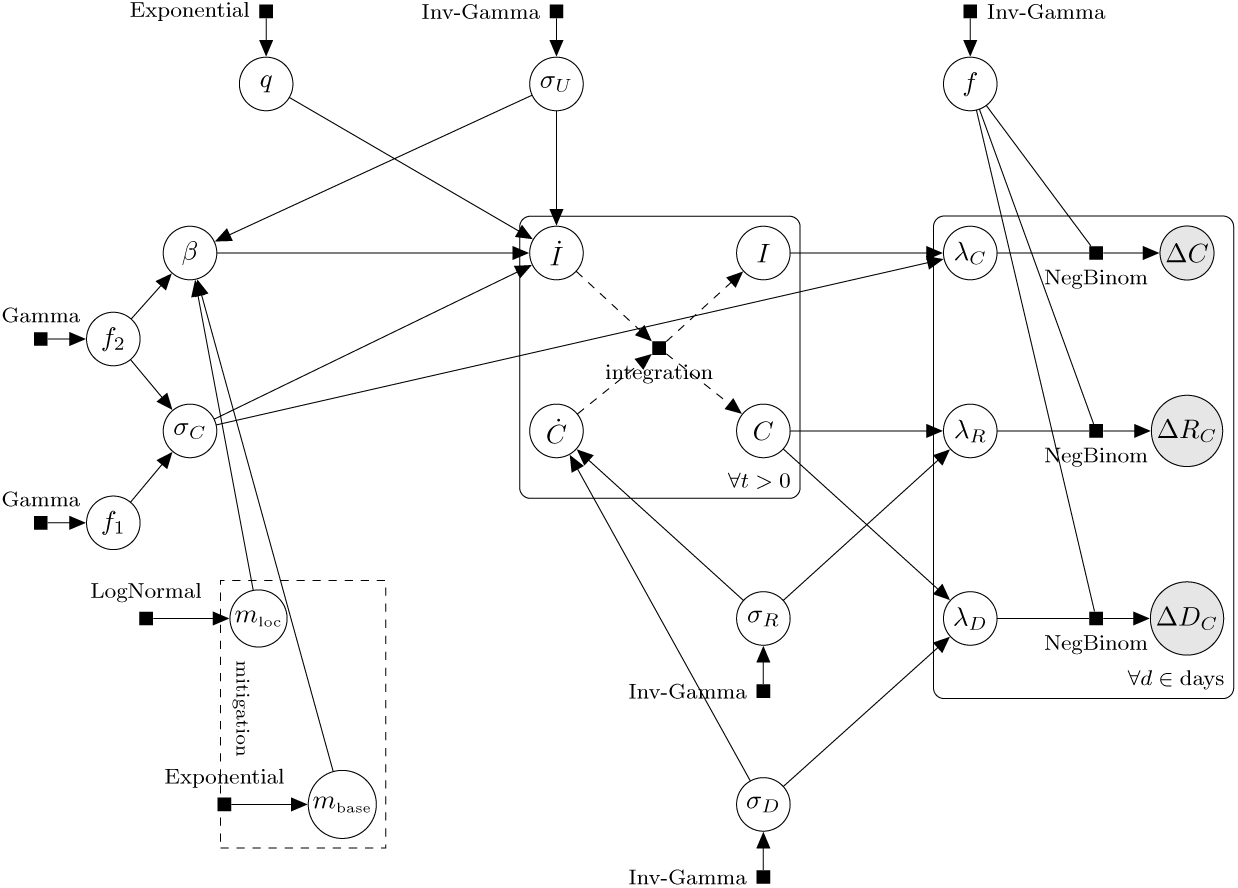
Bayesian Model diagram showing variable dependencies and prior distribution families.

The cumulative population that becomes infected and cumulative cases at time *t* are given by

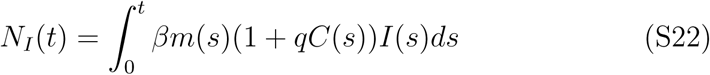

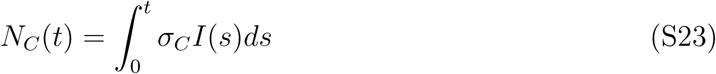

The cumulative totals for recovered and dead are *R_C_* and *D_C_*. From these quantities we can estimate the case ascertainment ratio, *N_C_/N_I_* and the total infection fatality ratio, *D_C_/N_I_*.

## S4 Data and Software

All data and code can be found at https://github.com/nih-niddk-mbs/covid-sicr. This repository contains several python scripts and Jupyter notebooks for replicating our findings, as described in the README file there. These can be used to obtain new post-publication estimates with additional data, provided that the data providers listed in the main text continue to provide a consistent API. We used Stan, which provides Bayesian inference using a heavily-optimized No-U-Turn sampler, a variant of Hamiltonian Monte Carlo, and each model described here is available as a .stan file. Python 3 was used for all other reported results, including interaction with Stan (via pystan). A subset of the work implemented in Julia is also available in the repository.

